# Rare Loss-of-function Mutations of *PTGIR* Identified in Fibromuscular Dysplasia and Spontaneous Coronary Artery Dissection

**DOI:** 10.1101/19012484

**Authors:** Adrien Georges, Juliette Albuisson, Takiy Berrandou, Délia Dupré, Aurélien Lorthioir, Valentina D’Escamard, Antonio F Di Narzo, Daniella Kadian-Dodov, Jeffrey W Olin, Ewa Warchol-Celinska, Aleksander Prejbisz, Andrzej Januszewicz, Patrick Bruneval, Anna A. Baranowska, Tom R. Webb, Stephen E. Hamby, Nilesh J. Samani, David Adlam, Natalia Fendrikova-Mahlay, Stanley Hazen, Yu Wang, Min-Lee Yang, Kristina Hunker, Nicolas Combaret, Pascal Motreff, Antoine Chédid, Béatrice Fiquet, Pierre-François Plouin, Elie Mousseaux, Arshid Azarine, Laurence Amar, Michel Azizi, Heather L. Gornik, Santhi K. Ganesh, Jason C. Kovacic, Xavier Jeunemaitre, Nabila Bouatia-Naji

## Abstract

**Background:** Fibromuscular Dysplasia (FMD) and Spontaneous Coronary Artery Dissection (SCAD) are related, non-atherosclerotic arterial diseases mainly affecting middle-aged women. Little is known about their physiopathological mechanisms.

**Objectives:** We aimed to identify rare genetic causes to elucidate molecular mechanisms implicated in FMD and SCAD.

**Methods:** We analyzed 29 exomes that included familial and sporadic FMD. Follow-up was conducted by targeted or Sanger sequencing (1,071 FMD and 365 SCAD patients) or lookups in exome (264 FMD) or genome sequences (488 SCAD), all independent and unrelated. We used TRAPD burden test to test for enrichment in patients compared to gnomAD controls. The biological effects of variants on receptor signaling and protein expression were characterized using transient overexpression in human cells.

**Results:** We identified one rare loss-of-function variant (LoF) (MAF_gnomAD_=0.000075) shared by two FMD sisters in the prostaglandin I2 receptor (hIP) gene (*PTGIR*), a key player in vascular remodeling. Follow-up in >1,300 FMD patients revealed four additional LoF allele carriers and a putative enrichment in FMD (P_TRAPD_=8×10^−4^), in addition to several rare missense variants. We confirmed the LoFs (Q163X and P17RfsX6) and one missense (L67P) to severely impair hIP function *in vitro*. Genetic analyses of *PTGIR* in SCAD revealed one patient who carries Q163X, one with L67P and one carrying a rare splicing mutation (c.768+1C>G), but not a significant enrichment (P_TRAPD_=0.12) in SCAD.

**Conclusions:** Our study shows that rare genetic mutations in *PTGIR* are enriched among FMD patients and found in SCAD patients, suggesting a role for prostacyclin signaling in non-atherosclerotic stenosis and dissection.

**Condensed abstract:** Fibromuscular Dysplasia (FMD) and Spontaneous Coronary Artery Dissection (SCAD) are non-atherosclerotic arterial diseases predominantly affecting women. Their mechanisms and genetic causes are poorly understood. We identified rare loss-of-function mutations of the prostacyclin receptor gene (*PTGIR*) in several FMD and SCAD patients, including two affected sisters, and several unrelated patients. We also showed that a rare missense mutation of *PTGIR* severely impairs prostacyclin receptor function *in vitro*. Our data provide evidence for a role for prostacyclin signaling in the etiology of FMD and SCAD providing leads towards this mechanism.

## Introduction

Fibromuscular Dysplasia (FMD) is an atypical and challenging vascular disease. FMD causes non-atheromatous stenosis, dissection, tortuosity and aneurysms in medium-sized arteries, mainly renal and carotid, but virtually all arteries can be affected (1). Although frequently asymptomatic, FMD is often associated with debilitating conditions such as hypertension and stroke and presents primarily in middle-aged women (80-90%), without hyperlipidemia or obesity (2).

The pathology and molecular mechanisms of FMD are poorly elucidated, with data coming mainly from observational studies using imaging and histology. Two main types of FMD lesions are defined using angiographic classification (1). The most common is multifocal FMD, which results in a “string-of-beads” appearance of the affected artery and represents more than 80% of cases. Focal FMD accounts for the rest of cases and is characterized by isolated stenosis. From the histology, the most commonly described lesions are medial fibroplasia, corresponding to the multifocal angiographic appearance, with an observed excess in fibrous connective tissue in the media of diseased arteries (1). An overall disorganization of the medial layer is also observed, with clear cellular loss of smooth muscle cells (SMCs) (3,4). The high proportion of early-middle age women among patients suggests a role for female hormones and hormone-associated vascular remodeling in the disease, although clear and direct mechanisms are still missing (5). Excessive pulsatility of arteries that results in the accumulation of micro-traumas was also proposed as a potential cause (6). Another suggested hypothesis is the occlusion of *vasa vasorum* that would result in intramural ischemia and myofibroblast transformation of SMCs (7).

Observational studies report a substantial clinical association between FMD and Spontaneous Coronary Artery Dissection (SCAD), an increasingly recognized cause of acute myocardial infarction in young to middle-aged women (8,9), with 25 to 86% of SCAD patients presenting FMD lesions in an additional arterial bed outside of the coronary circulation (8). FMD and SCAD share several clinical features; in particular, they both primarily affect middle-aged women and have a notable lack of classical atherosclerotic risk factors. SCAD is characterized by the obstruction of a coronary artery due to the presence of an intramural hematoma and/or a dissected intimal layer. As in the case of FMD, the pathological origin of SCAD lesions is unclear (8). An important additional aspect of the overlap between these diseases is that non-coronary arterial dissection (i.e. of the carotid arteries) is a common feature in FMD.

The absence of prospective epidemiological studies and animal models restricts our understanding of the natural history of or FMD and SCAD. A recent systems biology study suggested an association between *CD2AP* encoding CD2-associated protein and its plasma protein levels with FMD (10). The investigation of the genetic causes offers alternative angles to understand the molecular pathology and mechanisms behind arterial lesions. In a recent study, we identified the first genetic risk locus for FMD, a common variant located in the phosphatase and actin regulator 1 gene (*PHACTR1*) (11). We also found that the same risk allele for FMD was also at increased risk of SCAD, independently from the presence of FMD lesions among SCAD patients (12). This genetic link between FMD and SCAD supported a complex genetic pattern of inheritance for both diseases. Thus, a combination of genetic and environmental factors (local micro-trauma, hormonal fluctuation) could trigger diverse biological mechanisms that result in arterial remodeling and/or coronary artery dissection.

In a complex genetic model, rare pathogenic mutations may also represent causal factors with incomplete penetrance in some patients (13), as is the case for several cardiovascular diseases (e.g hypercholesteremia, coronary artery disease). To test this hypothesis, we applied exome sequencing to 29 individuals including FMD siblings and sporadic early onset cases to search for mutated genes with potential relevance to non-atherosclerotic arterial stenosis. We followed up one candidate gene in 1,335 FMD and 852 SCAD patients from four countries. We provide evidence for the prostacyclin receptor to harbor recurrent rare mutations in FMD and SCAD which result in loss of function of receptor signaling *in vitro*.

## Methods

A more detailed description of methods can be found in the supplementary appendix.

### Clinical origin of patients and diagnosis criteria

#### FMD patients

Familial FMD cases were ascertained as patients with at least one first-degree relative with confirmed FMD and were followed at the RVDRC. Sporadic FMD cases were recruited from the Rare Vascular Diseases Reference Center (RVDRC) of the European Hospital Georges Pompidou in Paris, France (HEGP), French ARCADIA (Assessment of Renal and Cervical Artery DysplasIA) registry, Polish ARCADIA-POL registry, US DEFINE-FMD study, University of Michigan Genetic Study of Arterial Dysplasia, Cleveland Clinic FMD Biorepository, as described previously (10-12,14). In all studies, an FMD diagnosis was established by clinical experts based on the observation of typical FMD-related lesions of middle-size arteries on imaging (computed tomographic angiography, magnetic resonance angiography, catheter-based angiography or duplex ultrasound in specialized centers) in absence of features of other causes of arterial stenosis such as atherosclerosis or vasculitis, biochemical evidence of inflammation, or syndromic arteriopathy. FMD experts reviewed imaging to independently confirm the diagnosis. Except for the exome-sequencing analysis, only unrelated cases were analyzed, as confirmed by the examination of medical records and array-based genotyping data, when available.

#### SCAD patients

SCAD patients were recruited from the French DISCO French register study, the UK SCAD Study, and the Victor Chang Cardiovascular Center (Australia). The diagnosis of SCAD was confirmed by review of the index coronary angiogram by an experienced interventional cardiologist with expertise in the recognition of SCAD, along with contemporaneous medical records. Individuals without a diagnostic angiogram were excluded from all genetic analyses. The majority of patients were of European origin. We obtained individual written informed consent from all participants included. Clinical characteristics of patients are presented in Table 1.

**Table 1:**
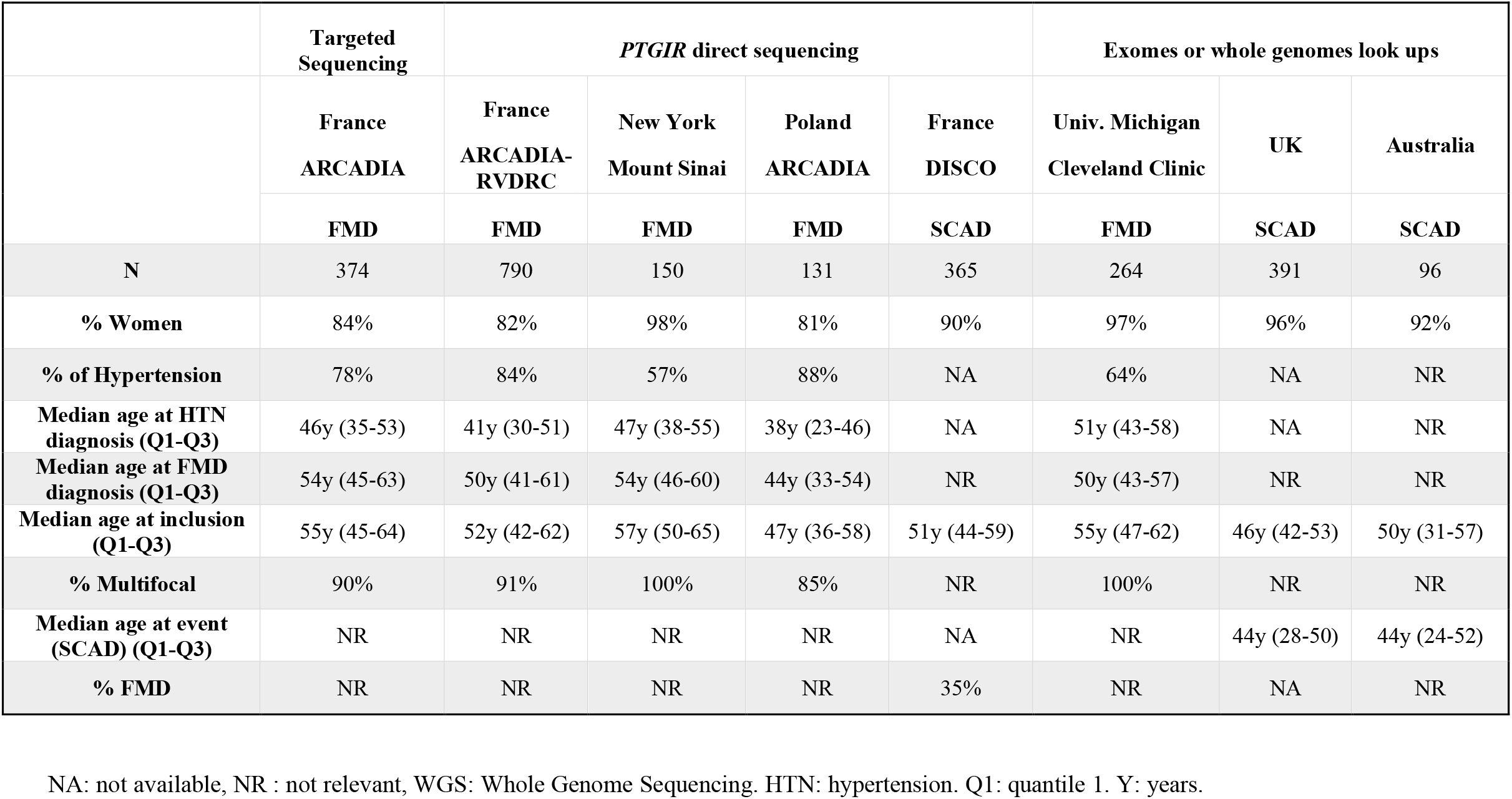
General Characteristics of Study Populations.

|  | Targeted Sequencing | *PTGIR* direct sequencing |  |  |  | Exomes or whole genomes look ups |  |  |
| --- | --- | --- | --- | --- | --- | --- | --- | --- |
|  | France<br>ARCADIA | France<br>ARCADIA-<br>RVDRC | New York<br>Mount Sinai | Poland<br>ARCADIA | France<br>DISCO | Univ. Michigan<br>Cleveland Clinic | UK | Australia |
|  | FMD | FMD | FMD | FMD | SCAD | FMD | SCAD | SCAD |
| N | 374 | 790 | 150 | 131 | 365 | 264 | 391 | 96 |
| % Women | 84% | 82% | 98% | 81% | 90% | 97% | 96% | 92% |
| % of Hypertension | 78% | 84% | 57% | 88% | NA | 64% | NA | NR |
| Median age at HTN diagnosis (Q1-Q3) | 46y (35-53) | 41y (30-51) | 47y (38-55) | 38y (23-46) | NA | 51y (43-58) | NA | NR |
| Median age at FMD diagnosis (Q1-Q3) | 54y (45-63) | 50y (41-61) | 54y (46-60) | 44y (33-54) | NR | 50y (43-57) | NR | NR |
| Median age at inclusion (Q1-Q3) | 55y (45-64) | 52y (42-62) | 57y (50-65) | 47y (36-58) | 51y (44-59) | 55y (47-62) | 46y (42-53) | 50y (31-57) |
| % Multifocal | 90% | 91% | 100% | 85% | NR | 100% | NR | NR |
| Median age at event (SCAD) (Q1-Q3) | NR | NR | NR | NR | NA | NR | 44y (28-50) | 44y (24-52) |
| % FMD | NR | NR | NR | NR | 35% | NR | NA | NR |

NA: not available, NR : not relevant, WGS: Whole Genome Sequencing. HTN: hypertension. Q1: quantile 1. Y: years.

#### Immunohistochemistry

For immunohistochemistry, paraffin blocks of arterial tissues (3 renal arteries) were obtained from surgical pathology archives of the European Hospital Georges Pompidou and belonged to patients who agreed to donate remaining tissue for research purposes. We used two normal renal arteries and one FMD artery where incident diagnosis was made from the histology.

### Study patients

Figure 1 summarizes the exome filtering and the follow-up strategies applied in this study.

**Figure 1:**
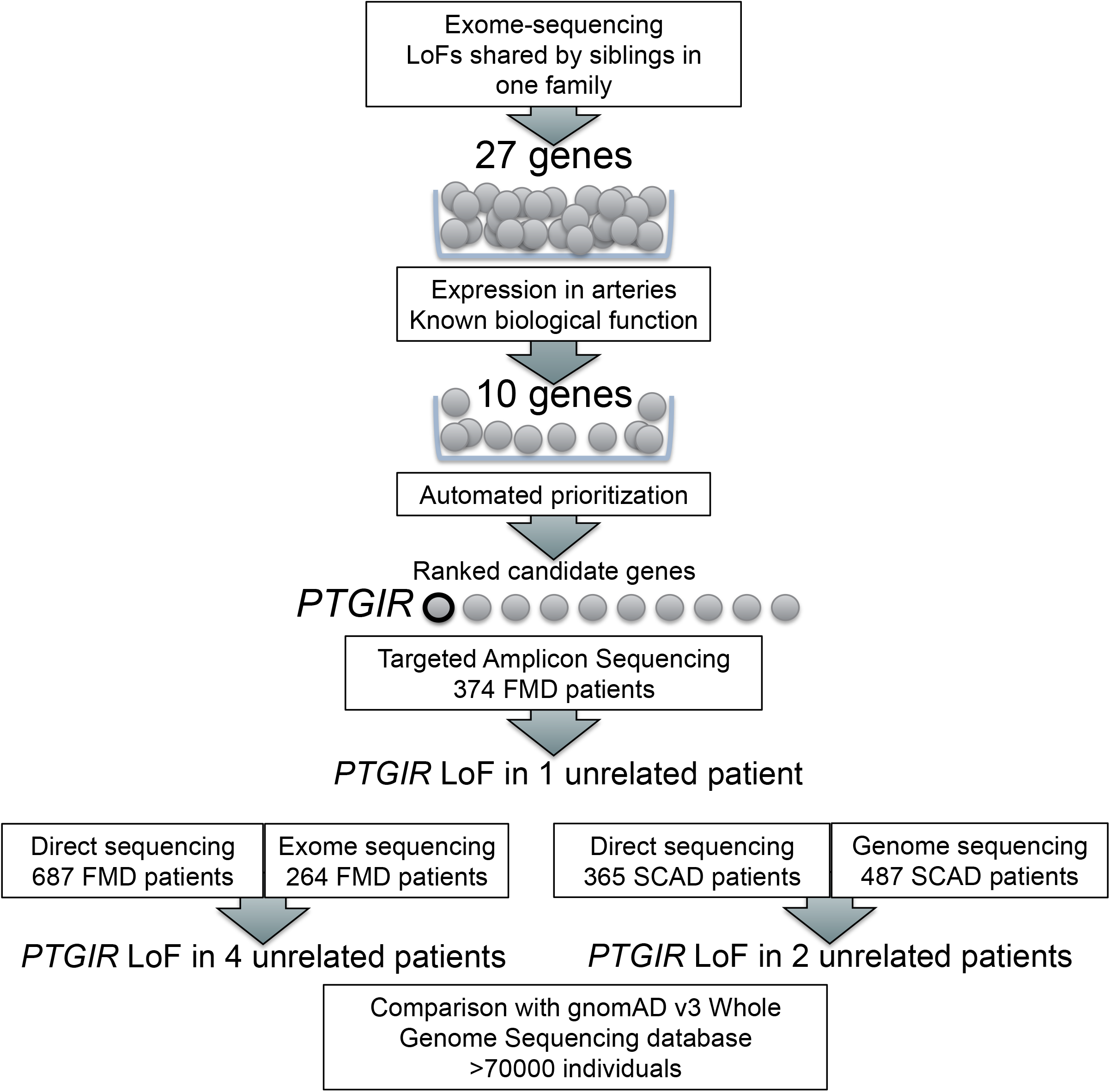
Flowchart illustrating the genetic approach undertaken in this study.

#### Exome sequencing patients

We studied 21 FMD familial cases formed by 4 FMD sib-pairs, 2 sib-trios, 1 cousins-pair and 1 family with 5 affected sibs (Supplementary Figure S1 and (15)). We also studied 4 sporadic FMD cases and had access to DNAs from unaffected parents of two of these cases, who were analyzed as family trios and had an early onset FMD (12 and 16 years old at diagnosis). In total, we analyzed 29 exome sequences. Exome and targeted re-sequencing were performed by Integragen® Genomics (Evry, France), as previously described (15).

We also interrogated whole exome sequences for mutations in *PTGIR* from 264 FMD patients (University Michigan), and whole genome sequences of 391 British (Leicester University) and 96 Australian (Victor Chang Cardiovascular Center) SCAD patients. Sanger sequencing was used to validate genotypes presenting low coverage (<10 reads) in the exome sequencing data.

#### Direct re-sequencing patients

Direct sequencing of *PTGIR* was performed in a total of 1,071 FMD patients. Seven hundred ninety were French patients from ARCADIA or the RVDRC, including the 374 analyzed by target resequencing, 150 were US patients from the DEFINE-FMD study and 131 were Polish from the ARCADIA-POL registry. We also screened 365 SCAD patients from the DISCO registry (12). Sanger sequencing was performed on a 3730xl DNA Analyzer system (Applied Biosystems).

### Molecular characterization of mutants

Wild-type or mutant hIP protein were overexpressed in human embryonic kidney cells (HEK293) for 48h. Cyclic adenosine mono-phosphate (cAMP) quantification was performed using cAMP-Glo assay (Promega, WI, USA), according to manufacturer’s description. Western Blot and immunofluorescence assays were performed as previously described (11). Mouse anti-hIP (sc-365268, Santa Cruz Biotechnologies, TX, USA) was used to detect prostacyclin receptor.

### Statistical analyses

Associations between *PTGIR* (LoF variants), FMD and SCAD were tested using a gene-based burden test implemented in TRAPD (Testing Rare vAriants using Public Data) package. We took advantage of the availability of latest version of gnomAD (v3) to evaluate the enrichment of rare *PTGIR* alleles in patients (16). GnomAD v3 uses only whole genome sequencing from >70000 individuals, which provides a homogeneous and large dataset with high read depth (>99.9% of samples with >15X coverage at PTG1R locus) allowing robust variant calling and estimation of the frequencies of these rare alleles. We used two-sided Fisher’s exact test to estimate the enrichment p-values, as recommended by the authors (16).

Unless otherwise noted, p-values were evaluated from a Student’s t-test in functional characterization experiments.

## Results

### Gene prioritization from exome sequencing data

We re-analyzed our previous exome sequences that included four affected sib pairs, two affected sib trios, and one cousin pair (15) in combination with a new sample of one sib quintet, two sporadic cases analyzed with both parents (two trios) and two unrelated sporadic cases (Figure 1, Figure S1). Relevant variants were defined as non-synonymous predicted deleterious or LoF unobserved or with low frequency in gnomAD (MAF<0.001). The inter-family analysis and trio filtrations were inconclusive with no gene containing relevant variants shared in at least two families, or recessive transmitted from parents to cases or *de novo* in the trios.

We then restricted the analyses to genes with LoF variants following the potential transmission pattern of each family, including dominant, recessive and compound heterozygotes. A short-list of 27 genes was identified to which we applied three prioritization tools (17-19). Using DAVID and STRING algorithms for functional annotations, we found that 10 genes had potential roles in blood vessel biology and signaling. We then provided a training list of genes involved directly or as regulators / effectors in stenosis and aneurysm related phenotypes in human diseases and mouse knockout models to the ENDEAVOUR tool. Using basic machine learning techniques to model the arterial dysfunctions observed in FMD, ENDEAVOUR ranked the prostaglandin receptor gene (*PTGIR*) as the best candidate. *PTGIR* contained one rare LoF (rs199560500 / Q163X, MAF = 0.00075 in gnomAD) that was shared by two affected sisters from Family 2. *PTGIR* encodes prostaglandin I2 (prostacyclin) receptor (hIP), a G protein-coupled receptor well known for its cardioprotective, anti-atherosclerotic and anti-thrombotic functions (20). For follow-up, we selected a short list of candidate genes based on prioritization tools, literature link with arterial stenosis, smooth muscle contraction or extracellular matrix organization, in addition to *PTGIR* (Table S1).

### Direct sequencing identifies recurrent LoF variants in *PTGIR* among FMD patients

We analyzed 20 genes using targeted amplicon sequencing in 374 patients (Table 1). A full list of identified LoF variants is shown in Table S1. We found truncating variants in five genes (*P2RX6, P2RY4, P2RY11 P2RX4*, and *PTGIR*). The variants identified in *P2RX6, P2RY4, P2RY11* and *P2RX4* were either as common in FMD patients as in gnomAD or direct sequencing did not confirm their presence. However, we found an additional patient carrying Q163X, the same LoF variant identified by exome sequencing in one FMD sib pair, which supported further the investigation of *PTGIR* coding sequences in more patients.

Overall, we analyzed 1,071 unrelated FMD patients by direct sequencing (Figure S2). First, we confirmed Q163X carriers among the 374 patients and screened both *PTGIR* coding exons in this sample, especially exon 2, which is incompletely covered by targeted amplicon sequencing. Then, we sanger sequenced 632 additional FMD patients from France (N=416), Poland (N=131) and US (N=150). We identified Q163X in one additional FMD patient and one rare frameshift variant in two FMD patients (rs754755149 / P17RfsX6, MAF=9×10^−5^ in gnomAD) (Table 2). We also looked up exome sequencing results in 264 FMD patients from the University of Michigan/Cleveland Clinic biorepository and found one additional carrier of Q163X at the heterozygote state. As the region coverage by exome sequencing at this locus was low in this cohort (3X depth), we confirmed the presence of the variant using Sanger sequencing (Figure S2).

**Table 2:** Description of rare loss-of-functions and missense variants identified in *PTGIR* in FMD patients.

| rs | cDNA | Protein | Consequence | FR | POL | NY | MI | gnomAD<br>(alleles/10000) |  | CADD<br>score | SIFT<br>score | Polyphen<br>score |
| --- | --- | --- | --- | --- | --- | --- | --- | --- | --- | --- | --- | --- |
|  |  |  |  | 790 | 131 | 150 | 264 | NFE | all |  |  |  |
| **rs754755149** | c.48del | P17RfsX6 | Frameshift | 2 | 0 | 0 | 0 | 1.2 | 0.9 | NA | NA | NA |
| **rs199560500** | c.487C>T | Q163X | Stop gained | 3 | 0 | 0 | 1 | 1.5 | 0.8 | 48 | NA | NA |
| **rs201261904** | c.4G>A | A2T | Missense | 1 | 0 | 0 | 0 | 0.5 | 1.0 | 16.9 | 0.1 | 0.1 |
| **rs1397542892** | c.200T>C | L67P | Missense | 0 | 0 | 1 | 0 | 0.2 | 0.1 | 31 | 0 | 1 |
| **rs775137134** | c.319A>G | M107V | Missense | 0 | 1 | 0 | 0 | 0.0 | 0.1 | 23.9 | 0 | 0.3 |
| **rs199969416** | c.409C>T | R137C | Missense | 1 | 0 | 0 | 0 | 0.0 | 0.0 | 24.9 | 0.1 | 0.9 |

LoF: loss-of-function variant. NA: not available, NFE: Non-Finnish Europeans. The number of alleles is indicated (no homozygous patients identified)

Using TRAPD burden test, we found that the FMD cohort of patients (N=1,335) was significantly enriched for LoF alleles in *PTGIR* compared to a control population from gnomAD v3 (N=71,702, P=4×10^−4^, TRAPD burden test, Table S2). All LoF carriers are of European ancestry. Similar enrichment was observed when we restricted the comparison to Non-Finnish European controls (N=32,399, P=8×10^−4^, Table S2).

### Functional characterization of rare LoF and missense variants in *PTGIR*

In addition to the two *bonafide* LoFs, we identified four rare missense variants in FMD patients (MAFgnomAD<0.001, Table 2) for which functional characterization is not reported. rs201261904 affects the N-terminal cytoplasmic extension of hIP (A2T), whereas the three other variants are located in the transmembrane helices (L67P, M107V, R137C, Figure S3a). Among these, rs1397542892 (L67P) affects a strictly conserved residue among vertebrate orthologs of *PTGIR* and is predicted to be deleterious (SIFT score of 0, Polyphen score of 1, Table 2, Figure S3b).

As mentioned, *PTGIR* encodes for the prostacyclin receptor. Therefore, to assess the potential impact of *PTGIR* rare variants on cellular functions, we overexpressed wild type and mutant hIP in the human embryonic kidney cell line HEK293 and measured production of cAMP in response to Iloprost, a synthetic analogue of prostacyclin. We measured a 50% maximal effective concentration of Iloprost (EC50_Ilo_) of 0.05nM for wild-type hIP (Figure 2). As expected, transfection with plasmids encoding any of the two nonsense mutants Q163X and P17RfsX6 resulted in a total loss of cell sensitivity to Iloprost (Figure 2a). All four missense mutant proteins were at least partially functional in overexpressing cells (Figure 2b). However, L67P hIP had a 100-fold decrease in Iloprost sensitivity (EC50_Ilo_ = 14.1nM, p=1.1 × 10^−4^, Figure 2c) whereas other mutants were not significantly different from wild-type hIP. Western Blot using anti-hIP antibody could detect all tested missense mutants, and the band pattern was similar between wild-type and mutant proteins (Figure 3a, Figure S4a), showing that no gross changes of post-translational modifications were caused by the mutations. L67P mutant expression was strongly affected, sometimes undetectable (Figure 3a), with an average signal reduction by 70 to 90% (N=4, p=0.03), whereas other mutants did not cause a significant reduction of hIP expression. Using immunofluorescence in HEK293 cells, we observed that wild-type hIP was mostly visible in the cytoplasm and at the plasma membrane (Figure 3b, Figure S4b). Conversely, we observed a clear colocalization of L67P hIP protein with intracellular organelles bound by Wheat Germ Agglutinin (WGA), a widely used lectin that binds to sialic acid and N-acetylglucosaminyl modified proteins, thus suggesting L67P mutation partly impairs hIP maturation (Figure 3b, Figure S5b). Finally, co-expression of wild-type hIP (fused with mCherry) with wild type or mutant versions of hIP did not affect cellular response to Iloprost (Figure S5a), which excludes the existence of a full dominant negative effect of hIP mutants. However, co-expressing hIP-mCherry protein with mutant hIP, we found that mature forms of hIP-mCherry were almost undetectable in presence of Q163X and L67P hIP, whereas the other mutants had no effect on hIP (Figure S5b). This suggests a potential dominant effect of these two mutants, providing a potential mechanism for the pathogenesis of heterozygous *PTGIR* mutations.

**Figure 2:**
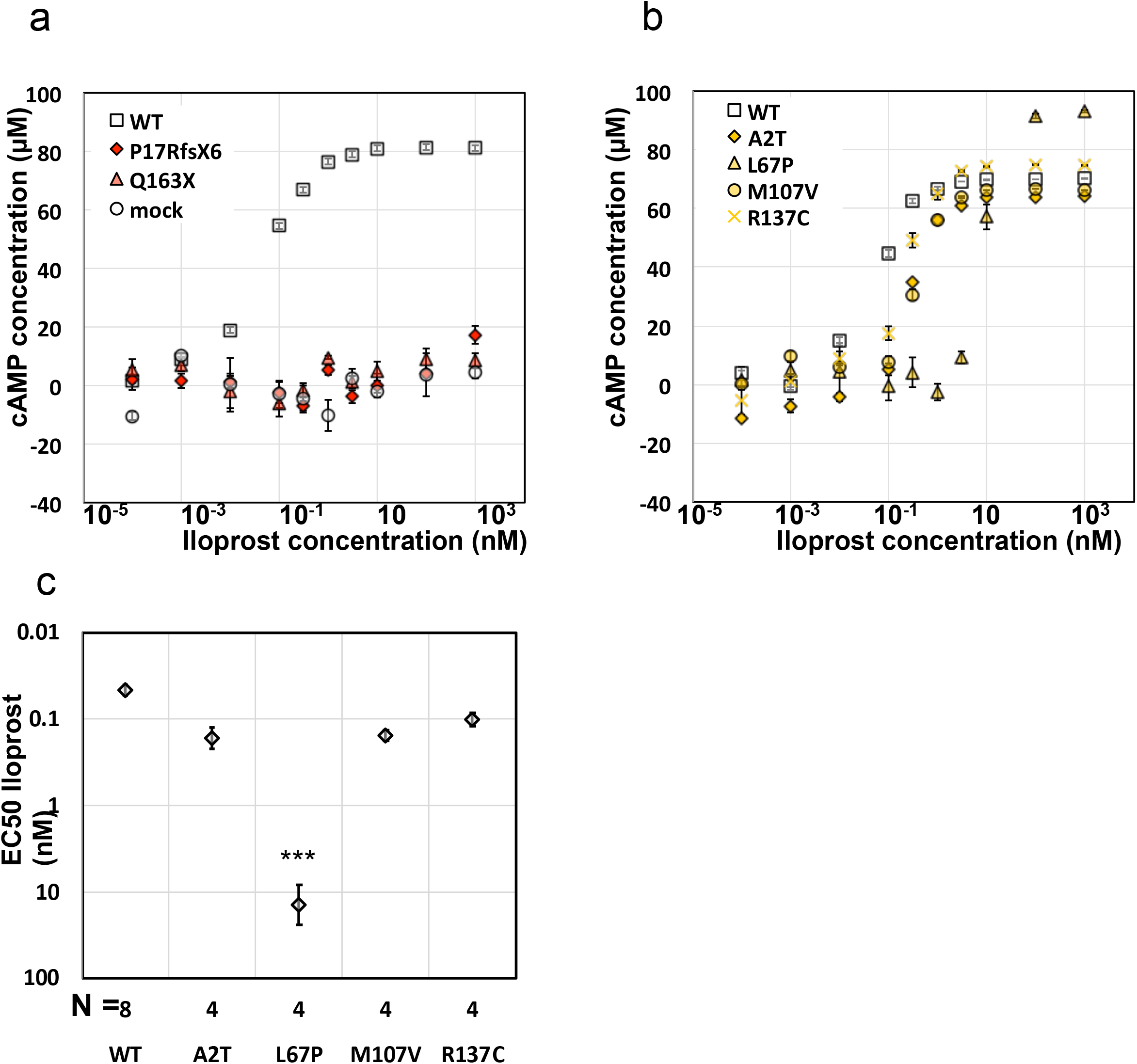
Analysis of cyclic adenosine monophosphate synthesis in response to iloprost in HEK293 cells overexpressing wild-type or mutant prostacyclin receptors. a-b) Concentration of cyclic adenosine monophosphate (cAMP) in HEK293 cells overexpressing wild-type (WT) prostacyclin receptor (hIP), mutant hIP (a : P17RfsX6 or Q163X, b: A2T, L67P, M107V or R137C) or transfected with mock plasmid (pcDNA-FLAG-HA)). Error bars represent the standard deviation of three biological replicates. c) Measurement of 50% response concentration to iloprost (EC50) in HEK293 cells overexpressing wild-type or mutant hIP. N represents the number of independent experiments. Error bars represent the standard error of the mean (SEM). Student’s t-test p-value (bilateral test with homoscedastic variance) ***: p < 10^−3^.

**Figure 3:**
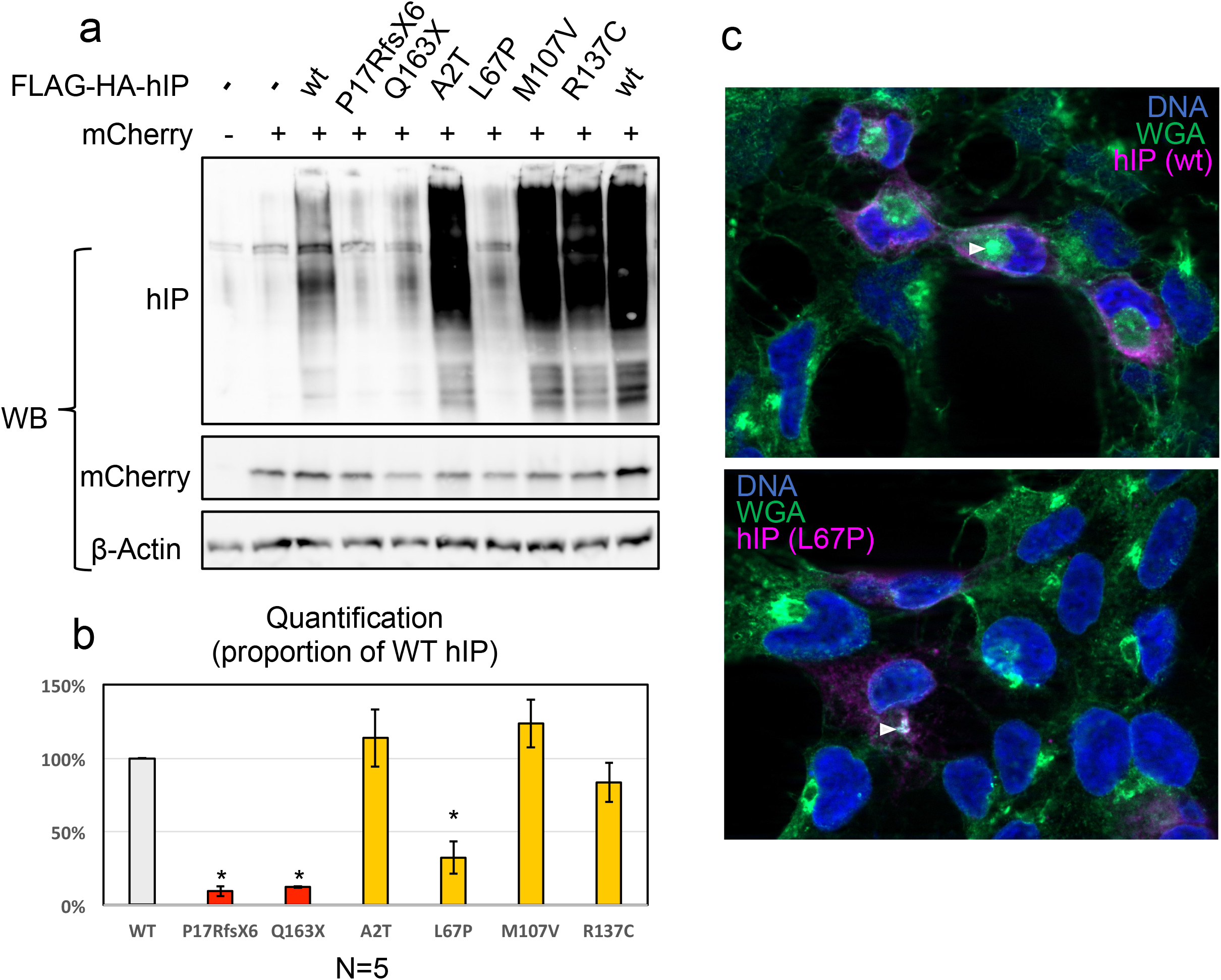
Evaluation of the expression and localization of the prostacyclin receptor mutated proteins. a) *Wild type and mutant protein expression*. SDS-PAGE/Western Blot assay on whole cells extracts of HEK293 cells overexpressing wild-type or mutant prostacyclin receptor (hIP) included a FLAG-HA N-terminal tag and the transfection control (mCherry). Proteins (hIP, mCherry and β-Actin) were detected using specific primary antibodies. We found that hIP is detected as a mix of bands and higher molecular weight smear due to its extensive post-translational modification. b) *Protein quantification*. We performed five independent experiments, and the average is shown with SEM ase error bars. We indicate Student’s t-test p-value (paired test)when p<0.05 as *. c) *Immunofluorescence visualization of HEK293 cells overexpressing wild type or L67P hIP*. Protein localization was assayed using hIP specific antibody (purple signal). Fixed cells were incubated with Alexa488-conjugated Wheat Germ Agglutinin (WGA, green signal), and with DAPI (blue signal). Images were taken with a 100x objective on a Zeiss ApoTome system.

### Clinical features of FMD patients carrying *PTGIR* non-functional alleles

The patients harboring *PTGIR* non-functional alleles were all women, with ages at FMD diagnosis ranging from 26 to 61 years (Table 3). Six of the seven patients had multifocal FMD, and one patient had only a focal lesion. Lesions were first identified in the renal artery for four patients and in the carotid artery for the other three patients. Five patients were screened for multivessel FMD, and two of them had FMD lesions in other vascular beds. Five patients had hypertension, with onset 1 to 10 years prior to FMD diagnosis, all of them having renal FMD. No dissection or aneurysm was detected in any of the seven patients. Patient 5, carrying the rs754755149 frameshift variant, was a 26-year-old woman with focal FMD affecting one renal artery. She underwent angioplasty, which was initially successful, but restenosis was seen during follow-up. All other patients had multifocal lesions. Patient 7, carrying L67P missense, underwent angioplasty of both renal arteries, restoring normal blood flow with a moderate benefit on the management of hypertension. Upon follow-up for >5 years, no restenosis was detected. The other patients had no angioplasty.

**Table 3:** Clinical characteristics of FMD patients carrying non-functional *PTGIR* alleles.

|  | Patient 1 | Patient 2 | Patient 3 | Patient 4 | Patient 5 | Patient 6 | Patient 7 |
| --- | --- | --- | --- | --- | --- | --- | --- |
| *rs ID* | rs199560500 |  |  |  | rs754755149 |  | rs1397542892 |
| cDNA position | c.487C->T |  |  |  | c.48del |  | c.200T->C |
| Protein position | Q163X |  |  |  | P17RfsX6 |  | L67P |
| Sex (M/F) | F | F | F | F | F | F | F |
| Age at inclusion (years) | 61 | 53 | 46 | 57 | 26 | 62 | 65 |
| Age at diagnosis (years) | 61 | 52 | 46 | 56 | 26 | 61 | 62 |
| Familial (Y/N) | Y | N | N | ND | N | N | N |
| FMD subtype | Multifocal | Multifocal | Multifocal | Multifocal | Focal | Multifocal | Multifocal |
| Multisite (Y/N/ND) | ND | N | N | Y | ND | N | Y |
| Number of vascular beds | ND | 1 | 1 | 3 | ND | 1 | 3 |
| Arterial beds involved | Renal (both) | Carotid (both) | Renal (unilateral) | Carotid, iliac, renal | Renal (unilateral) | Renal (both) | Internal carotid, vertebral, renal |
| Hypertension (Y/N) | Y | N | Y | N | Y | Y | Y |
| Age at onset (years) | 51 |  | 46 |  | 24 | 61 | 62 |
| Dissection ? (Y/N) | N | N | N | N | N | N | N |
| Known aneurysms ? (Y/N) | N | N | N | N | N | N | N |
| Required intervention (dilation) ? (Y/N) | N | N | N | N | Y (+restenosis) | ND | Y (no restenosis) |
| Smoking status | Smoker | Former smoker | Smoker | Smoker | Smoker | Non-smoker | Non-smoker |

Patients are organized in three groups, according to the rare variant they carry. Y: yes, N: no, ND: not determined

### Histological localization of the prostacyclin receptor

Immunochemistry staining of three renal arteries from pathology archives (two normal, one FMD) with human prostacyclin receptor (hIP) antibody showed protein presence in all arterial layers, with a stronger expression in SMCs of the medial layer, both in normal and FMD tissues (Figure S6). Of note, none of the patients carrying *PTGIR* mutations had vascular surgery, which limited access to their arteries to evaluate the mutant proteins expressions.

### Rare LoF mutations also identified in SCAD patients

Given the established clinical and genetic overlap between FMD and SCAD, we also screened 365 French SCAD patients from the DISCO registry for rare variants in *PTGIR* by direct sequencing. We found one patient with a splicing variant not reported in gnomAD (rs1302581755). The variant is a G to C substitution in position +1 of the 3’ end of exon 2, disrupting the donor splicing site, resulting in a predicted LoF (Table S3). We also found one more patient with L67P mutation. A look up for variants in genome sequences of 391 British and 96 Australian SCAD patients identified one additional patient carrying the Q163X, rs199560500 variant allele (Table S3). We also identified additional rare missense variations of *PTGIR* in SCAD patients, for which functional effects are not known (Table S3). Although *PTGIR* LoF alleles were relatively more frequent in SCAD patients than in gnomAD control populations, we did not find a significant enrichment for LoFs in *PTGIR* among SCAD patients compared to gnomAD controls using TRAPD burden test (P = 0.12).

### Clinical features of SCAD patients carrying *PTGIR* LoF alleles

The SCAD patients were all women, aged 38 to 66 years at first SCAD event (Table 4). SCAD patient 1, bearing rs199560500 variant, presented with non-ST elevation myocardial infarction, hypertension on treatment with a history of gestational hypertension. Upon coronary angiography, a very extensive dissection of the left mainstem coronary could be identified, with no signs of atherosclerosis. The two other patients were older at the time of their SCAD event (58 and 66 years), and SCAD lesions presented as a diffuse stenosis leading to a partial or complete occlusion of the coronary artery, with a visible hematoma in the arterial wall. Upon full screening of other arterial beds, these three SCAD patients did not exhibit detectable FMD lesions.

**Table 4:** Clinical characteristics of SCAD patients carrying non-functional *PTGIR* alleles.

|  | **Patient 8** | **Patient 9** | **Patient 10** |
| --- | --- | --- | --- |
| **rs** | rs199560500 | rs1397542892 | rs1302581755 |
| **cDNA** | c.487C->T | c.200T->C | c.768+1C>G |
| **Protein** | Q163X | L67P | ND |
| **Sex (M/F)** | F | F | F |
| **Age at 1st event (years)** | 38 | 66 | 58 |
| **Clinical presentation** | Non-STEMI | STEMI | Non-STEMI |
| **SCAD subtype** | 1 | 2 | 2 |
| **P-SCAD (Y/N)** | N | N | N |
| **Recurrent SCAD (Y/N)** | N | N | N |
| **Other ischemic events** | N | Y | N |
| **Migraine (self-reported)** | N | Y | N |
| **Hypertension (Y/N)** | Y | N | N |
| **Dyslipidemia (Y/N)** | N | N | N |
| **Smoking status** | ND | Non-smoker | Smoker |
| **FMD in other arterial beds (Y/N)** | N | N | N |

Y:yes, N: no, ND: not determined. SCAD subtype is based on Saw classification.

## Discussion

Here we describe rare loss-of-function and missense mutations in the prostacyclin receptor gene (*PTGIR*) in FMD and SCAD patients (Central Illustration). We found that these mutations severely impair prostacyclin receptor signaling *in vitro* and are prevalent at a rate significantly higher in a cohort of ∼1,300 FMD patients, compared to large unselected publicly available cohorts. We also describe some of these rare mutations in SCAD patients, without an overt significant enrichment. Our study describes an unprecedented genetic impairment in the prostacyclin signaling in non-atheromatous arterial stenosis and dissection.

### Further support for a shared genetic basis between FMD and SCAD

Understanding the accurate genetic model has been a challenging endeavor for FMD and SCAD. We have recently established their complex genetic mode of inheritance through the association between the common variant rs9349379 in *PHACTR1* and an increased risk for both diseases (11,12). After a first negative exome sequencing study in FMD (15), we increased the sample numbers and applied a prioritization strategy that allows the identification of *PTGIR* as a novel gene mutated in FMD. A recent exome study conducted on SCAD families described rare mutations in *TLN1* (21). The identification of *PTGIR* as an additional gene involved in FMD and potentially SCAD further supports their complex genetic basis involving both common and rare mutations. We suspect incomplete penetrance for the mutations described in *PTGIR*. Several LoF variants are reported in gnomAD, which include population-based and diseased cohorts, but in lower frequency compared to FMD patients. On the other hand, the high estimated prevalence of asymptomatic FMD in unselected populations (1-6%) (1) is compatible with a substantial number of undiagnosed FMD patients carrying *PTGIR* mutations among these large public cohorts. Here we estimate in our cohorts that at least 0.5% of FMD patients carry a non-functional *PTGIR* allele. Of note, this rate of *PTGIR* mutation carriers is comparable to the percentage reported for *PSCK9* mutation carriers (0.6%) among cohorts of hypercholesterolemia (22).

### Rare non-functional LoFs in *PTGIR* among FMD and SCAD patients

We provide evidence for complete loss of protein function *in vitro* caused by the two LoFs and showed that missense mutations are functional, especially the L67P that strongly impairs the receptor signaling and protein function. Previous studies have implicated *PTGIR* missense variants, in particular rs4987262 (R212C), in atherosclerosis and thrombosis (23). It should be noted that this variant is relatively frequent (1% in Non-Finnish European populations in gnomAD) and was identified in 22 FMD patients and 12 SCAD patients in our cohorts, including one SCAD patient with homozygous mutation (Table S3). Apart from L67P mutation, all missense mutations studied here show a 2 to 5-fold decrease in sensitivity to Iloprost, similar to previous observations for R212C mutation (24). We note that several rare missense variants identified in SCAD patients only were not functionally characterized. It is therefore possible that we underestimate the pathogenicity of *PTGIR* missense mutations.

### The prostacyclin signaling and physiopathology of arterial stenosis and dissection

The loss or decrease in prostacyclin signaling has a wide range of biological consequences, all compatible with arterial remodeling and dissection observed in FMD and SCAD patients. Prostacyclin is a well-established vasodilator, with anti-thrombotic and anti-atherosclerotic properties. Prostacyclin is also a potent repressor of the proliferation and migration of SMCs from the vascular wall and promotes their contractile phenotype (20), which is compatible with the lesions observed in FMD. A high level of fibrotic tissue characterizes FMD lesions. Interestingly, prostacyclin signaling is known to repress fibrosis in several tissues (20). The cAMP signaling downstream hIP is a well-known regulator of fibroblast function and a repressor of fibrosis through the direct inhibition of ECM synthesis, myofibroblast differentiation and fibroblast proliferation (25). Prostacyclin effects are often seen as opposite to the effects of Thromboxane A2, a closely related prostaglandin, which favors thrombosis and vasoconstriction, and their balance is a key to several pathologies (20). Aspirin, widely prescribed for its anti-thrombotic properties, inhibits the production of both prostacyclin and thromboxane and may compensate the effect of unbalanced prostacyclin/thromboxane signaling in FMD patients. However, anti-thrombotic medication may represent an additional risk in patients developing aneurysms in middle-size arteries, and the screening for *PTGIR* mutations may help identify patients that could benefit of such medication. On the other hand, Iloprost is used to treat pulmonary arterial hypertension, scleroderma, Raynaud’s phenomenon and other diseases with prominent vasoconstriction, and may have unpredictable effects in patients bearing hIP mutations (20). Whether FMD and SCAD patients with *PTGIR* mutations may exhibit increased platelet aggregation and an increased risk of developing pulmonary arterial hypertension or thrombosis would be an interesting investigation to conduct in the future. Given the high relevance of the prostacyclin to thromboxane balance to vascular function, the genetic investigation of more genes linked to these pathways could provide additional clues about the role of this mechanism in FMD and SCAD.

### Study limitations

The prevalence of the identified *PTGIR* LoFs is low and affects a small fraction of FMD and SCAD patients. On the other hand, the study samples are relatively limited, compared to other complex cardiovascular diseases and may result in inaccurate estimation in mutation rate among patients. We lack large pedigrees with clinical and genetic information in affected and unaffected members to assess the extent of the penetrance of *PTGIR* mutations. We do not provide significant support for enrichment in SCAD, compared to FMD, potentially due to differences in samples sizes. We identified heterozygous LoFs, with biological consequences that are hard to predict considering the multiple biological functions of hIP, and its possible interactions/compensations with related prostaglandin receptors in a more complex physiological system.

## Conclusions

We identified genetic defaults in *PTGIR* that impair its cellular function and are likely to be rare genetic causes for FMD and SCAD. According to our genetic screen of FMD and SCAD cohorts, we estimate *PTGIR* mutations to be present in ∼0.5% of FMD patients and ∼0.3% of SCAD patients. Larger studies are needed to refine these estimations, which should include a greater number of patients. This finding, and the availability of multiple drugs targeting this pathway, may help clinicians to design specific therapeutic approaches for FMD and SCAD patients. Further genetic analyses involving functionally related genes are required to fully evaluate the influence of this pathway on the pathogenesis of non-inflammatory stenosis and dissection observed in FMD and SCAD arteries.

## Clinical perspectives

This study adds evidence to the possibility of FMD and SCAD share a common genetic basis. We show that rare loss of function variants in the gene encoding the prostacyclin receptor (*PTGIR*) are enriched in FMD patients and present in SCAD patients. This pathway is a target of widely used drugs such as aspirin or iloprost. If this mechanism is confirmed by further larger genetic and clinical studies, these findings may help the clinicians identify the best therapeutic strategy to treat FMD and SCAD patients in the future.

## Data Availability

Data reported are individual mutations describes in a small sample of patients and are described in detail and made available in the manuscript. All raw data of the in vitro experiments is available to be shared if requested.

## Acknowledgements

We thank Drs Robert Graham and Eleni Giannoulatou from Victor Chang Cardiac Research Institute, Sydney, New South Wales, Australia for providing exome-sequencing data for SCAD patients. We thank recruiting clinicians in all centers and the patients who participated in this study. We acknowledge the Fibromuscular Dysplasia Society of America for enabling study enrollments at meetings into the University of Michigan Genetic Study of Arterial Dysplasia. The authors thank AstraZeneca’s Centre for Genomics Research, Discovery Sciences, BioPharmaceuticals R&D for funding the sequencing of and providing the bioinformatics support related to subjects recruited at Leicester University. We acknowledge the leadership of the ESC-ACCA SCAD Study Group and thank our patients and volunteers for their support.

## Abbreviations

FMD: Fibromuscular dysplasia
SCAD: Spontaneous Coronary Artery Dissection
LoF: loss-of-function mutation
hIP: human prostacyclin receptor
*PTGIR*: prostacyclin I2 receptor gene
TP: Thromboxane A2 Receptor
cAMP: cyclic adenosine mono-phosphate
SMC: smooth muscle cell

---

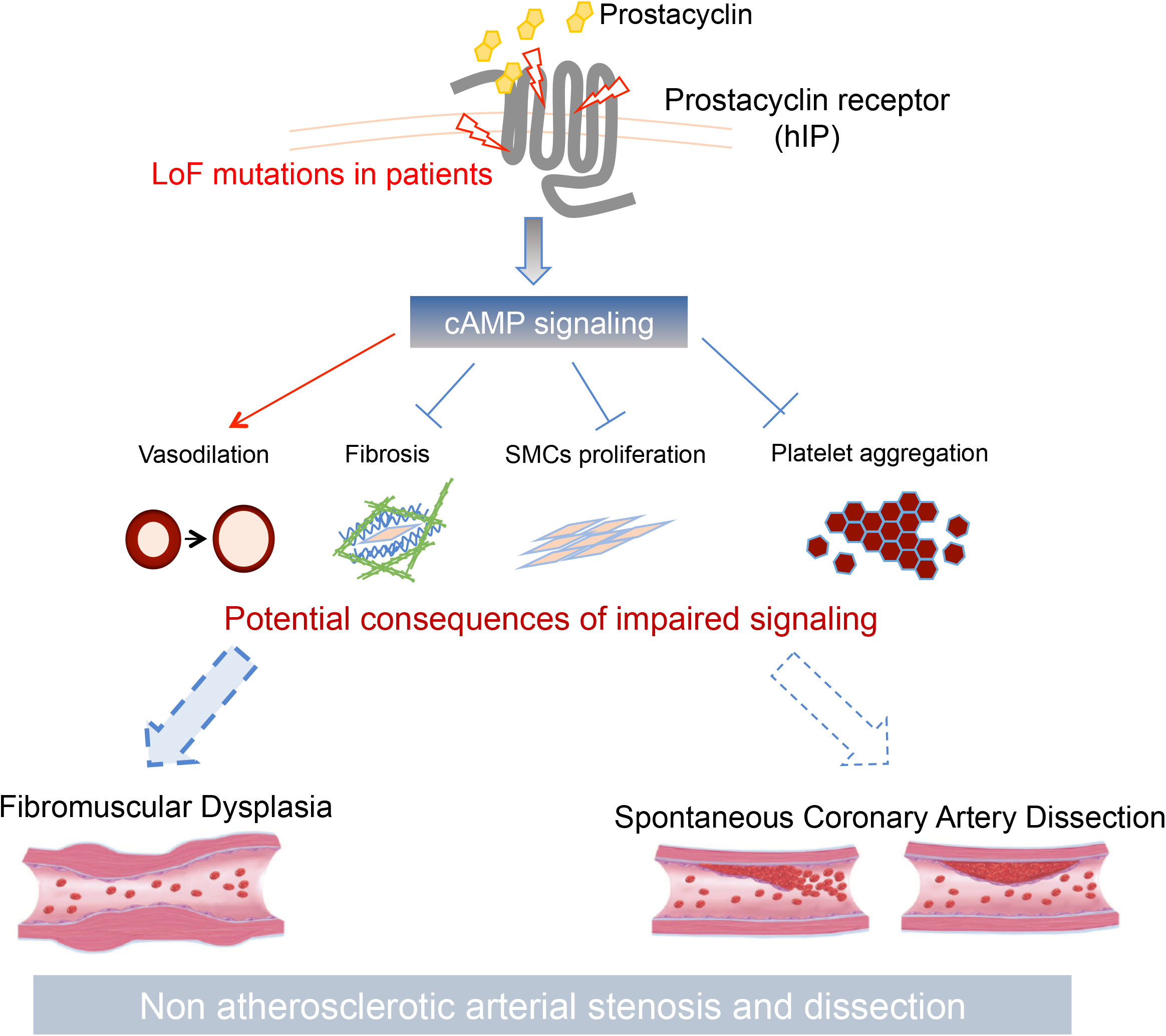

## Supplemental material

### Methods

#### Familial cases

FMD cases were recruited from the French database of the Rare Vascular Diseases Referral Centre of the European Hospital Georges Pompidou (HEGP), Paris, France. An expert panel using clinical information from the medical history, the examination and the interview of the patient, the interpretation of angiography and/or computed tomography scan defined the FMD diagnosis after the exclusion of other causes of arterial stenosis (mainly atherosclerosis, Takaysu disease and Elhers Danlos syndrome). Familial FMD cases were ascertained as patients with at least one first-degree relative with confirmed FMD. We identified eight pedigrees from which we selected at least two patients per pedigree with DNA quality and quantity compatible with exome sequencing experiments. Selected patients are sibs in all families, except in Family 7 wherein they are first cousins (Figure S1). Intrafamily controls could not be identified, as angiography explorations in healthy members of these families were not available. We also selected 2 early onset cases (<20 years at diagnosis) for whom both unaffected parents could be contacted and were willing to participate in the study, and 2 severe sporadic cases for whom parents could not be contacted. The study was approved by local institutional ethics committee (Approval CCPPRB Paris-Cochin #RBM 00-028) and all individuals gave written informed consent.

#### ARCADIA-RVDRC

We analyzed unrelated FMD followed-up at the Rare Vascular Diseases Reference Center (RVDRC) of the European Hospital Georges Pompidou (HEGP), Paris, France. We ascertained patients from the ARCADIA (Assessment of Renal and Cervical Artery DysplasIA) register, an ongoing national FMD registry at the HEGP, Paris. The diagnosis of FMD in RVDRC and ARCADIA patients was established using clinical information from the medical history, the interpretation of angiography and/or computed tomography scan of arterial beds after the exclusion of other causes of arterial stenosis such as atherosclerosis, Takayasu disease and Elhers Danlos syndrome and neurofibromatosis type 1. Given the complexity of the interpretation of imaging of vascular diseases, a local panel of experts including clinicians from the departments of hypertension, radiology, vascular medicine and medical genetics validated the diagnosis of FMD. French FMD study protocol was approved by the Ile-De-France research ethics committee (Comité de Protection des Personnes: CPP d’île de France) on 03/04/2009 (ID: 2009-A00288-49). In the context of ARCADIA registry, women and men aged ≥18 years, diagnosed with RA or eCVA FMD, were prospectively recruited at 16 university hospitals in France and Belgium. Most participating centers were approved by the European Society of Hypertension as Hypertension Excellence Centers or by the French Ministry of Health as FMD Competence Centers. Specialists in hypertension, vascular medicine, vascular neurology, and vascular radiology were available in all centers. The protocol was approved by the Comité de Protection des Personnes Ile-de France II. All participants provided written informed consent. The procedures followed were in accordance with the institutional guidelines.

#### ARCADIA-POL

All patients were women and men aged over 18 years entering a Polish multicenter registry involving 32 centers in Poland with coordination of Department of Hypertension Institute of Cardiology. Each of these centers followed referral pattern and identified patients with newly diagnosed or established or suspected renal FMD, FMD in any vascular bed, or spontaneous artery dissection (particularly in carotid, vertebral, or coronary arteries). Only FMD patients were screened in this study.

All patients underwent detailed clinical evaluation including ambulatory BP measurements (ABPM), biochemical evaluation, biobanking, duplex Doppler of carotid and abdominal arteries. In all patients, a computed tomography angiography of intracranial and cervical arteries as well as a computed tomography angiography of abdominal aorta and its branches including coeliac trunk, mesenteric arteries, common hepatic artery, splenic artery, renal arteries and iliac arteries were performed during one hospital stay. FMD was diagnosed as nonatherosclerotic arterial encroachment or stenosis affecting the trunk or branches of medium size arteries, in the absence of aortic wall thickening, biochemical evidence of inflammation and known syndrome arterial disease. Renal FMD was divided into multifocal and unifocal types are preciously defined. Unifocal FMD corresponded to a single stenosis on a given vessel, regardless of its length, and multifocal FMD to at least two stenoses on a given vessel segment. Patients with FMD lesions affecting at least two of the four predefined vascular beds (renal artery, extracranial carotid and vertebral arteries, intracranial arteries, mesenteric/splenic arteries and iliac arteries) were classified as having multisite FMD, and the others (irrespective of the presence of unilateral or bilateral FMD lesions of the paired arteries) were classified as single-site FMD. The diagnosis of FMD and its types was made by two independent investigators. The ARCADIA-Pol protocol was approved by local authorities (authorization number : IK-NP-0021-101/1482/16).

#### The DEFINE-FMD Study

DEFINE-FMD is a systems biology study aiming to DEFINE key disease drivers and mediators of FMD. Its forerunner pilot study, the CAUSE study (ClinicalTrials.gov Identifier: NCT01808729), enrolled its first ‘run-in’ control subject on 31 October 2012, and first FMD patient on 22 February 2013. As the CAUSE study approached target enrolment it was closed (at n = 34 subjects) and we initiated DEFINE-FMD (ClinicalTrials.gov Identifier: NCT01967511). Both the CAUSE and DEFINE-FMD studies were approved by the institutional review board of the Icahn School of Medicine at Mount Sinai, and all subjects gave written informed consent. The investigation conformed to the principles outlined in the Declaration of Helsinki.

All FMD cases were seen and assessed in the Mount Sinai Vascular Medicine Clinic. Inclusion criteria for entry into DEFINE-FMD include ≥18 years of age, being freely willing to participate, and fluency in English. FMD cases are required to have a clinical diagnosis of multifocal FMD that is confirmed by imaging [computed tomographic angiography (CTA), magnetic resonance angiography, or catheter-based angiography]. While DEFINE-FMD was recently expanded and is now also enrolling subjects with spontaneous coronary artery dissection (SCAD) or cervical artery dissection (CvAD) in the absence of typical multifocal FMD, these subjects with isolated SCAD or CvAD were not included in this analysis. However, for this analysis confirmed multifocal FMD cases were permitted to have SCAD and/or CvAD. Exclusion criteria (for cases and controls) include: co-morbidities which reduce life expectancy to 1 year; any solid organ or hematological transplantation, or those in whom transplantation is considered; active autoimmune disease; illicit drug use; HIV positive; prior malignancy.

#### University of Michigan Genetic Study of Arterial Dysplasia and Cleveland Clinic FMD biorepository

Patients were recruited at the University of Michigan (UM) and/or the Cleveland Clinic. UM FMD cases were recruited into an IRB approved study through a referral clinic at the University of Michigan and through self-referral to the study (University of Michigan IRB number HUM00044507 and Cleveland Clinic FMD biorepository IRB number 10–318). The Cleveland Clinic cases were enrolled among consecutive patients seen at a dedicated FMD referral clinic. A vascular medicine specialist ascertained clinical diagnosis of FMD after review of diagnostic imaging and prior to blood sample collection. DNAs from healthy controls without vascular disease were obtained from the Cleveland Clinic Gene Bank, which was approved by the Cleveland Clinic IRB. Genomic DNA was isolated from a peripheral blood sample and analyzed as described (3).

#### The DISCO Study

French patients were recruited through the DISCO protocol, a nation-wide study aiming to assess the presence of FMD and its genetic determinants in a SCAD cohort using the clinical criteria defined in (1,2). Patients were included prospectively and retrospectively following inclusion criteria of age > 18 years, provided consent for genetic studies, and a confirmed diagnosis of SCAD by clinical presentation, angiography analyses confirmed by optical coherence tomography or intravascular ultrasound. (Clinical Trials ID: NCT02799186, approved by regional committee CPP Sud-Est 6 2016 AU-1258). DNAs from patients were genotyped by direct sequencing and were mostly Europeans.

#### The UK SCAD study

The UK SCAD study is ethically approved by the NHS Health Research Authority (14/EM/0056) subjects were recruited from the UK mainland. All participants were Europeans defined as such using genome-wide genotypes and/or clinical records.

#### Australia SCAD study

Patients from Australia were identified largely through a social media platform. The study was approved by the Human Research Ethics Committee of St. Vincent’s Hospital, Sydney. Genotyping was performed by Sanger sequencing (Garvan Molecular Genetics, Australia, NATA ISO17025 and ISO15189 certified). Controls were healthy subjects all >70 years old, available through the Medical Genome Reference Bank (MGRB) that involves genomic data performed by the Kinghorn Centre for Clinical Genomics, Australia. All participants were Europeans defined using genome-wide genotypes and/or clinical records.

#### Sequencing

##### Exome-sequencing and gene filtration strategy

Exome and targeted re-sequencing were performed by Integragen® Genomics (Evry, France), as previously described (15). Briefly, 3µg genomic DNA was sonicated to 150-200bp, and paired-end Illumina were ligated on repaired, A-tailed fragments. Purified, and PCR-amplified material was hybridized to Sureselect oligo probe library for 24h and affinity-captured. Eluted fragments were further amplified and sequenced on an Illumina HiSeq 2000 instrument (paired-end 75bp run). Sequences were mapped to hg19 genome using ELANDv2 algorithm and variant calling were performed using Illumina pipeline (CASAVA1.8).

Analysis of variations identified by exome sequencing was made using PolyWeb, an in house software (Paris Descartes Platform) that allows filtering-out of irrelevant variants (common, synonymous, low coverage, and predicted by Polyphen2 or SIFT as benign). This tool allows the comparison of results of patients and their relatives and takes in account the dominant, recessive and de novo modes of inheritance. Family structures of individuals were indicated when available, the three isolated cases were considered as independent families. In silico characterization and prioritization of genes with LoFs shared by members of the same family was performed using DAVID, a web-based tool for functional annotation and clustering of list of genes, STRING, to find functional protein association networks and ENDEAVOUR, a method that allows prioritization of genes using basic machine learning techniques to model the biological process under study and then to score and rank the candidate genes using that model (4-6).

##### Targeted amplicon/Next-Generation Sequencing

Targeted enrichment was performed with a PCR method on the Access Array microfluidic support from Fluidigm. 320 primer pairs were designed with Primer3, for an average product size of 261pb. Library preparation followed the description of the Access Array User Guide with Integragen optimizations (AmplIG) with increased level of multiplexing. Forty eight pools of 332 PCR obtained from one Access Array were subjected to a second round of PCR for 10 cycles in a standard microplate format in order to add specific barcodes for sample identification and P5/P7 Illumina adapters for Illumina sequencing. The 48 pools were then controlled and quantified on Fragment Analyzer to perform 1 equimolar pool of 64 samples. Finally one run of one lane of MiSeq V2, 2×150b was launched for 64 samples.

Base calling was performed using the Real-Time Analysis software sequence pipeline (2.7.6) with default parameters. Sequence reads were mapped to the human genome build (hg19 / GRCh37) for 2X analysis and a fasta file with amplicons sequences for 1X analysis using Elandv2e (Illumina, CASAVA1.8.2) allowing multiseed and gapped alignments. CASAVA1.8.2 was used to call single-nucleotide variants (SNVs) and short insertions/deletions (max. size is 300nt), taking into account all reads per position. SNVs and indels with Q(SNPs) < 10 and Q(Indel) < 20, or regions with low mappability (QVCutoff < 90) were filtered out. Variants annotation was based on dbSNP (dbSNP144), the 1000 Genomes Project (phase1_release_v3.20101123), the Exome Variant Server (ESP6500SI-V2-SSA137), and the Exome Aggregation Consortium (ExAC r3.0) and an in-house databases. Functional consequences of variants on genes were predicted by Variant Effect Predictor (VEP release 83), using SIFT (sift5.2.2) and PolyPhen (2.2.2) scores in the case of missense mutations.

##### Sanger sequencing

*PTGIR* fragments (four amplicons) were amplified using primers containing identical Illumina tails (0.5µM each) with JpStart Taq ReadyMix (Sigma Aldrich) with the following program: initial denaturation at 95°C for 3 minutes, then 35 cycles of 20s at 95°C, 20s at 60°C, 20s at 72°C, and a final 7 minutes extension step at 72°C. Twenty ng genomic DNA were used per reaction. 10µL of the reaction were cleaned of remaining dNTPs and primers by incubating with 0.4 unit FastAP Thermosensitive Alkaline Phosphatase (Thermo Fisher Scientific) and phosphatase and 2 units Exonuclease I (Thermo Fisher Scientific) for 30 minutes at 37°C, and enzymes were deactivated at 86°C for 10 minutes. 2µL of the reaction mix (from 16 µL) was used for sequencing reaction using BigDye™ Terminator v3.1 Cycle Sequencing Kit (Applied Biosystems) according to manufacturer’s instructions. Illumina tail F primer was used for all sequencing reactions. Sequencing was performed on a 3730xl DNA Analyzer system (Applied Biosystems).

For Sanger sequencing performed on University of Michigan samples, sequencing was performed as follow. Primers were designed around rs199560500 location. 10ng of genomic DNA of case and control was amplified using Phusion HF taq (NEB, Ipswich MA) and the following primers: FOR 5’ CTTCGCCTTCGCCATGACCTTCTTC 3’, REV 5’ CTGCCCTCCTCCTCTCCCCACTCC 3’. A 534bp amplicon was isolated and gel purified (Takara Bio, Moutain View, CA) and sent for Sanger sequencing (University of Michigan, Advanced Genomics Core).

##### Whole exome sequencing of FMD cases from the University of Michigan cohort

From each research participant, either blood or saliva samples were obtained using standard K+ EDTA tubes or commercial collection kits (Oragene, DNAGenotek). DNA was isolated according to the protocol of commercial kits (Nucleospin Tissue (TakaraBio), prepIT-L2P (DNAGenotek) and quantified using the Quant-iT PicoGreen dsDNA kit (ThermoFisher). Samples were randomized onto 96-well barcoded plates for exome sequencing at the Northwest Genomics Center (NWGC, University of Washington, Seattle WA) and shipped to the NWGC sequencing center. Quality control and sample tracking throughout the WES pipeline was conducted via sex-typing and sample fingerprinting, conducted through high frequency genotyping of cosmopolitan single nucleotide variants informative for individual identity. Samples with adequate DNA concentration and integrity, accurate sex-typing, and high quality fingerprint assay were sequenced.

Library Production, Exome Capture, Sequencing: Library construction and exome capture were performed using an automated (Perkin-Elmer Janus II) 96-well plate format. 1 ug of genomic DNA was subjected to a series of shotgun library construction steps, including fragmentation through acoustic sonication (Covaris), end-polishing, A-tailing and ligation of sequencing adaptors with dual 8 bp barcodes for multiplexing, followed by PCR amplification. Libraries underwent exome capture using SeqCap EZ Exome v2.0 Target Enrichment Probes (∼36.5 MB target) (Roche/Nimblegen, Pleasanton, CA). Prior to sequencing, the library concentration was determined by fluorometric assay and molecular weight distributions verified on the Agilent Bioanalyzer (consistently 150 ± 15 bp).

Sequencing: Barcoded exome libraries were pooled using liquid handling robotics prior to clustering (Illumina cBot) and loading. Massively parallel sequencing-by-synthesis with fluorescently labeled, reversibly terminating nucleotides was carried out on the HiSeq sequencer, with 8 exomes multiplexed per lane.

Read Processing: NWGC’s sequencing pipeline is a combined suite of Illumina software and other industry standard software packages (i.e., Genome Analysis ToolKit [GATK], Picard, BWA-MEM, SAMTools, and in-house custom scripts). The pipeline includes base calling, alignment, local realignment, duplicate removal, quality recalibration, data merging, variant detection, genotyping and annotation. Variants in the PTGIR gene were extracted for review.

##### Variant analysis

Frequencies were retrieved from gnomAD v3 considering the Non-Finnish European (NFE) and general population (all) as potential references. Combined Annotation Dependent Depletion (CADD) score aggregates many different indicators including conservation scores, ENCODE data and protein level scores to predict potential deleterious effect of SNVs by evaluating the likelihood of these annotations. In the scaled metric shown here, a CADD score >30 means that the variant belongs to the 0.1% of variants for which annotations are the less likely to appear by chance, indicating possible deleterious effect of the variation. Sorting Intolerant from Tolerant (SIFT) and Polymorphism Phenotyping (PolyPhen) scores predict whether a given amino acid substitution may affect protein function based on sequence homology and physical properties of amino acids. A SIFT score close to 0 or a PolyPhen score close to 1 indicate a possible deleterious alteration. CADD, SIFT and PolyPhen scores were recovered from ENSEMBL variant webpage (www.ensembl.org).

#### Statistical analyses

To test association between PTGIR gene (LoF variants) FMD we applied a gene-based burden test, which compares the overall burden of rare protein-modifying variants in a given gene in case and control subjects, using large-scale public sequencing databases such as Genome Aggregation Database (gnomAD) as control subjects. This approach is implemented in TRAPD package (7). Assuming a dominant model for *PTGIR* gene, we tabulate the number of individuals who carry at least one qualifying variant in our case cohort (n=1335) For the gnomAD control dataset (n= 71702 for all samples, n= 32299 for Non-Finnish Europeans only), the number of control subjects carrying at least one qualifying variant in *PTGIR* was approximated by the sum of all the qualifying variants allele counts in that gene. Finally, a two by two contingency table was constructed from tabulation of case and control counts. Table S2 represents the distribution of FMD/SCAD cases and control subjects according to the presence of qualifying variant in *PTGIR*. We used a two-sided Fisher’s exact test to estimate the association p-values.

#### Cell culture and transfection

HEK293 cells were grown in Dulbecco’s Modified Eagle Medium (DMEM) containing 4.5g/L Glucose, Glutamax™ and sodium pyruvate (Gibco), supplemented with 10% Fetal Bovine Serum (Corning) and 1% Penicillin/Streptomycin (Gibco). For transfection, cells were seeded at 40% confluence and transfected using Fugene HD (Promega), according to manufacturer’s instructions. To prepare precipitates, Fugene and DNA were mixed in a 2.5 µLFugene/µgDNA ratio in culture medium and incubated for 5 minutes, before adding to the cells.

#### Plasmids preparation

A full list of primers and plasmids used in this study can be found in Tables S4 and S5. All PCR steps were performed using Phusion Flash High-Fidelity PCR Master Mix (Fisher Scientific, Hampton, NH, USA), according to manufacturer’s instructions. The complete sequence of human *PTGIR* was amplified from human genomic DNA with primers containing restriction enzyme sites and subcloned between NheI and HindIII sites of pcDNA-FLAG-HA plasmid (addgene #52535, (8)). To generate PTGIR expression plasmids, coding regions from exon2 and exon3 were amplified separately with an overlapping primer. The two products were mixed with DNA Polymerase, and missing ends were elongated. Primers containing restriction enzyme sites were then added to amplify the complete coding sequence of PTGIR, and the digested product was integrated between XhoI/BamHI sites of pcDNA-FLAG-HA. To generate point mutants, two fragments on each side of the mutation were amplified, using a common primer containing the mutated sequence. The two products were mixed with DNA Polymerase, missing ends were elongated, and the complete product, containing the mutation, was then amplified using external primers containing restriction enzyme sites and integrated into pcDNA-FLAG-HA as before. To generate a PTGIR-mCherry expression plasmid, the sequence of mCherry was amplified and inserted between NotI and BamHI sites of pcDNA-FLAG-HA. Then PTGIR sequence (without stop codon) was amplified and inserted between NheI and NotI sites, removing FLAG-HA tag at the same time. mCherry control expression plasmid was created by inserting mCherry between NheI and XhoI sites of pcDNA-FLAG-HA, also removing FLAG-HA tag.

#### Cyclic Adenosine mono-Phosphate (cAMP) quantification

HEK293 cells were seeded at 40% confluence in two wells of 6-wells plates, and transfected on the next day with PTGIR expression plasmids. 48h after transfection, cells were trypsinized and counted. Cells were seeded in white opaque 96-wells plates (Corning, NY, USA) at a density of 20000 cells/well. After 24h, cells were washed once with PBS and treated with 20µL induction buffer (Dulbecco’s PBS (Fisher Scientific, Hampton, NH, USA) containing 500µM isobutyl-1-methylxanthine (Sigma Aldrich, MS, USA) and 100µM 4-(3-butoxy-4-methoxybenzyl) imidazolidone (Sigma Aldrich, MS, USA), alone or containing 10^−13^ to 10^−6^ M Iloprost (Sigma Aldrich, MS, USA). After 25 minutes incubation, cell lysis and measure of cAMP concentration were performed using cAMP-Glo assay (Promega, WI, USA), according to manufacturer’s instructions.

#### Western Blot

HEK293 cells were seeded at 40% confluence in 12-wells plates, and transfected on the next day with PTGIR expression plasmids and/or mCherry expression plasmids. 48h after transfection, cells were washed once with PBS and 100µL 2X Laemmli Sample buffer (Biorad, CA, USA) was added to each well. Cells were scrapped and collected into 1.5mL tubes, then incubated at 95°C for 10 minutes, with 900rpm agitation. Lysates were then centrifuged for 5 minutes at 12000g, and supernatants were frozen at −20°C.

Frozen samples were then thawed, supplemented with 5µL β-Mercaptoethanol (Sigma Aldrich, MS, USA) and incubated at 95°C for 5 minutes. 15µL of lysates were loaded and separated onto Mini-PROTEAN 12% TGX precast gels (Biorad, CA, USA). Proteins were transfered to a Protran Nitrocellulose membrane (Whatman, Maidstone, UK). Membranes were incubated 1h in Tris-buffered Saline supplemented with 0.05% Tween (Sigma Aldrich, MS, USA) (TBST) and 5% non-fat milk. Membranes were then incubated with the following primary antibodies in TBST supplemented with 2% non-fat milk : mouse anti-hIP 1/500 (sc-365268, Santa Cruz Biotechnologies, TX, USA), mouse anti-β-Actin 1/1000 (sc-47778, Santa Cruz Biotechnologies, TX, USA), rabbit anti-mCherry 1/1000 (ab167453, Abcam, Cambridge, UK), mouse anti-FLAG M2 1/1000 (F1804, Sigma Aldrich, MS, USA). After three 10min washes, membranes were then incubated with 1/20000 HRP-anti-mouse-IgG or HRP-anti-rabbit-IgG for 1h. After three 10min washes, revelation was performed using SuperSignal West Pico or Femto Fast Western Blot Kit (Fisher Scientific, Hampton, NH, USA), and images taken using a LAS4000mini system (GE Healthcare, IL, USA). Image processing and quantification were performed using Fiji ImageJ software (fiji.sc).

#### Immunofluorescence staining

HEK293 cells were seeded at 30% confluence on 10mm glass coverslips in 24 wells plates, and transfected on the next day with PTGIR expression plasmids. 48h after transfection, cells were washed once with DPBS and fixed for 15 minutes with 4% Formaldehyde (Fisher Scientific, Hampton, NH, USA) in DPBS. Cells were then washed three times with DPBS and incubated 10 minutes with 5µg/mL Alexa Fluor 488-conjugated Wheat Germ Agglutinin (W11261, Fisher Scientific, Hampton, NH, USA). Cells were washed again 3 times in DPBS and permeabilized with 0.5% Triton X-100 in DPBS for 15 minutes. Cells were then blocked with DPBS+5% non-fat milk for 1h, then incubated for 2h with primary antibody (mouse anti-PTGIR (sc-365268, Santa Cruz Biotechnologies, TX, USA)) diluted to 1/100 in the same buffer. After 3 DPBS washed, cells were incubated for 1h with secondary antibody (Goat anti-Mouse-IgG Alexa 647 (A28181 Fisher Scientific, Hampton, NH, USA)) at a 1/500 in DPBS/5% milk. Cells were washed 3 times in DPBS, then incubated with 1µg/mL DAPI for 15 minutes, washed again 3 times in DPBS, and twice in ddH2O. Coverslips were mounted on microscope slides using Aqueous Mounting Medium (CTS011, R&D Biosystems, MN, USA), and imaged using a Zeiss Axioimager Z2 Apotome system (Zeiss, Oberkochen, Germany), with a 100x objective. Raw images were processed using Fiji ImageJ software (fiji.sc).

#### Immunohistochemistry staining

Protein staining were performed on paraffin embedded artery samples using anti-hIP (1/100, sc-365268, Santa Cruz Biotechnologies, TX, USA), anti-MYH11 (1/100, sc-6956, Santa Cruz Biotechnologies, TX, USA) and anti-α-Actin (1/100, α-Actin smooth muscle antibody (1/100, α-ASM, Clone1A4, Dako, Trappes, France) primary mouse antibodies and revealed using an ABC peroxidase kit with diaminobenzidine (Vector laboratories, Burlingame, CA, USA). The arterial tissues were fixed in formalin and embedded in paraffin. Antigen retrieval were performed by incubating tissue sections in alkaline solution (Dako, Trappes, France) for 40 minutes at 94°C in a hot water bath.

## Supplementary Tables

**Table S1:** LoFs identified by targeted amplicon sequencing. The following genes were analysed : *CREM, LPAR4, LPAR6, LTB4R, P2RX1, P2RX2, P2RX3, P2RX4, P2RX5, P2RX6, P2RX7, P2RY1, P2RY11, P2RY12, P2RY13, P2RY14, P2RY2, P2RY4, P2RY6, PHACTR1, PTGIR, YY1AP1*. ^£^ SNP not in gnomAD, but a frequency of 0.07 is

| *Gene* | *Var_name* | *Pos (hg19)* | *Nomenclature* | *Consequence* | *Codons* | *Codons_AA* | *CADD* | *gnomAD ALL* | *gnomAD NFE* | *N* | *HET* | *HOM* |
| --- | --- | --- | --- | --- | --- | --- | --- | --- | --- | --- | --- | --- |
| *P2RX4* | rs773993953 | 12:121648093 | c.126C>G | stop | TAC/TAG | Y42* | 37 | 0.00001 | 0 | 4 | 4 | 0 |
| *P2RX5* | rs746000416 | 17:3594282 | c.328delA | frameshift | aCC/CC | T110Pfs38* | - | NA<sup>£</sup> | NA<sup>£</sup> | 8 | 7 | 1 |
| *P2RX6* | rs148541070 | 22:21369548 | c.55A>T | stop | AAG/TAG | K19* | 36 | 0.00377 | 0.00629 | 1 | 1 | 0 |
| *P2RY11* | rs146974153 | 19:10225189 | c.900C>G | stop | TAC/TAG | Y300* | 36 | 0.00087 | 0.00012 | 2 | 2 | 0 |
| *P2RY4* | rs41310667 | X:69478432 | c.1043G>A | stop | TGG/TAG | W348* | 36 | 0.01201 | 0.0196 | 16 | 13 | 3 |
| *P2RY4* | rs148362984 | X:69479172 | c.303G>A | stop | TGG/TGA | W101* | 37 | 0.00085 | 0.00145 | 1 | 1 | 0 |
| *PTGIR* | rs199560500 | 19:47126996 | c.487C>T | stop | CAG/TAG | Q163* | 37 | 0.00009 | 0.00015 | 1 | 1 | 0 |

reported in the UK10K cohort (Ensembl). NA: not available

**Table S2:** List of loss of function variants TRAPD burden assay. HET: Heterozygous, HOM: Homozygous, DOM: Dominant, REC: Recessive. Bold: variants identified in FMD/SCAD patients.

| N FMD | gnomAD | N | Variants considered | Cases |  |  | Controls |  |  | P DOM |
| --- | --- | --- | --- | --- | --- | --- | --- | --- | --- | --- |
|  |  |  |  | HET | HOM | Total | HET | HOM | Total |  |
| 1335 | all | 71702 | **rs754755149**, rs931307567,<br>**rs199560500**, rs1302581755,<br>19:46623845:G:T,<br>19:46623903:AGCAT:A,<br>19:46623318:C:A,<br>19:46623829:GC:G,<br>rs761595464, 19:46623970:G:A | 6 | 0 | 6 | 48 | 0 | 48 | 4.51x10<sup>-4</sup> |
| 1335 | NFE | 32299 | **rs754755149**, **rs199560500**,<br>rs1302581755,<br>19:46623318:C:A,<br>19:46623829:GC:G,<br>rs761595464, 19:46623970:G:A | 6 | 0 | 6 | 23 | 0 | 23 | 8.37x10<sup>-4</sup> |
| N SCAD | gnomAD | N | Variants considered | Cases |  |  | Controls |  |  | P DOM |
|  |  |  |  | HET | HOM | Total | HET | HOM | Total |  |
| 852 | all | 71702 | rs754755149, rs931307567,<br>**rs199560500**, **rs1302581755**,<br>19:46623845:G:T,<br>19:46623903:AGCAT:A,<br>19:46623318:C:A,<br>19:46623829:GC:G,<br>rs761595464, 19:46623970:G:A | 2 | 0 | 2 | 48 | 0 | 48 | 0.12 |
| 852 | NFE | 32299 | rs754755149, **rs199560500**,<br>**rs1302581755**,<br>19:46623318:C:A,<br>19:46623829:GC:G,<br>rs761595464, 19:46623970:G:A | 2 | 0 | 2 | 23 | 0 | 23 | 0.13 |

**Table S3:**
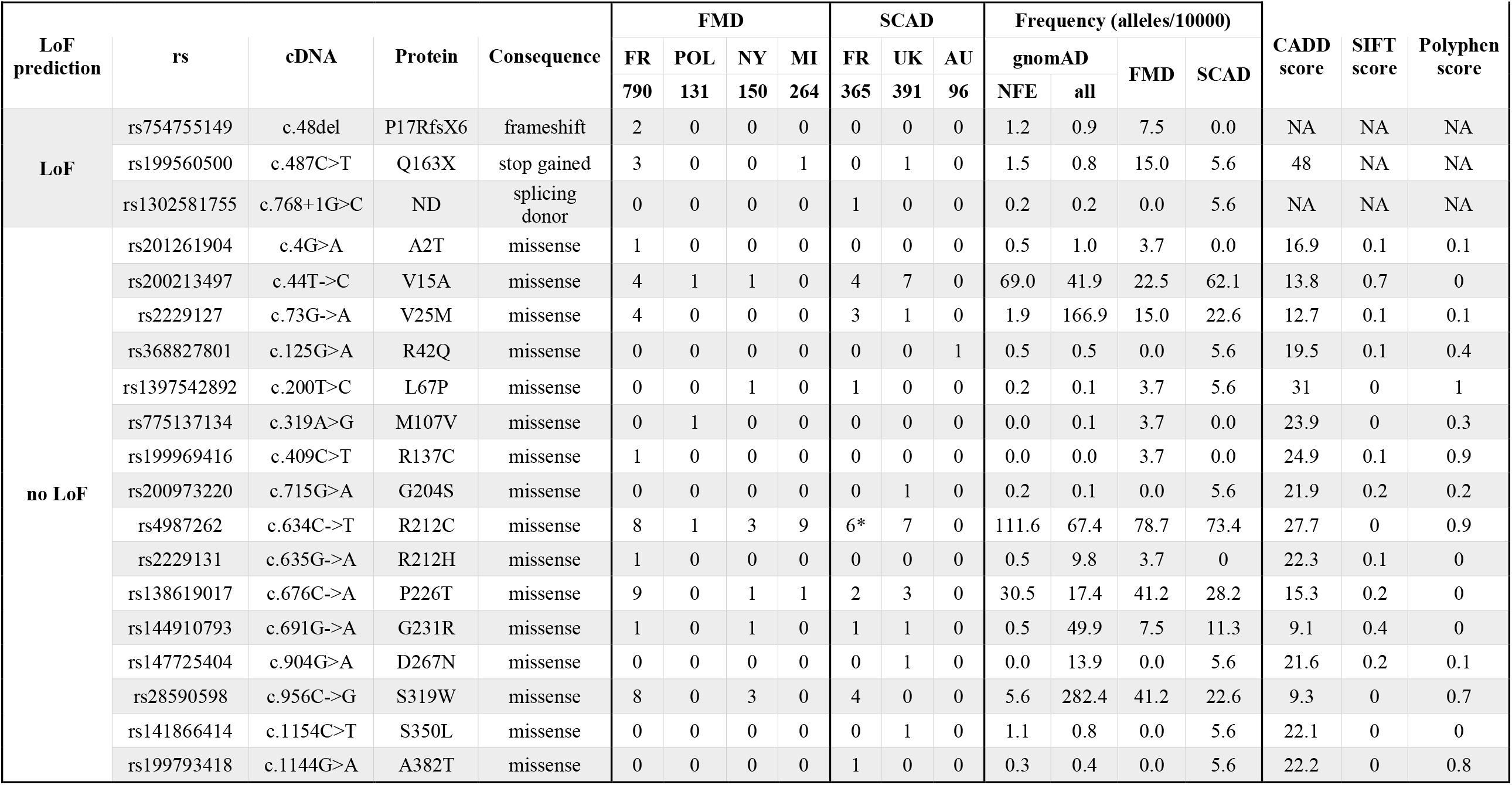
Description of all variants identified in PTGIR in FMD and SCAD patients (frequency <0.01 in gnomAD) The number of alleles is indicated. *: including one homozygous patient (5 patients with at least one allele). NA: not-available. ND: not determined. NFE : Non-Finnish Europeans

| LoF prediction | rs | cDNA | Protein | Consequence | FMD |  |  |  | SCAD |  |  | Frequency (alleles/10000) |  |  |  | CADD score | SIFT score | Polyphen score |
| --- | --- | --- | --- | --- | --- | --- | --- | --- | --- | --- | --- | --- | --- | --- | --- | --- | --- | --- |
|  |  |  |  |  | FR | POL | NY | MI | FR | UK | AU | gnomAD |  | FMD | SCAD |  |  |  |
|  |  |  |  |  | 790 | 131 | 150 | 264 | 365 | 391 | 96 | NFE | all |  |  |  |  |  |
| LoF | rs754755149 | c.48del | P17RfsX6 | frameshift | 2 | 0 | 0 | 0 | 0 | 0 | 0 | 1.2 | 0.9 | 7.5 | 0.0 | NA | NA | NA |
|  | rs199560500 | c.487C>T | Q163X | stop gained | 3 | 0 | 0 | 1 | 0 | 1 | 0 | 1.5 | 0.8 | 15.0 | 5.6 | 48 | NA | NA |
|  | rs1302581755 | c.768+1G>C | ND | splicing donor | 0 | 0 | 0 | 0 | 1 | 0 | 0 | 0.2 | 0.2 | 0.0 | 5.6 | NA | NA | NA |
| no LoF | rs201261904 | c.4G>A | A2T | missense | 1 | 0 | 0 | 0 | 0 | 0 | 0 | 0.5 | 1.0 | 3.7 | 0.0 | 16.9 | 0.1 | 0.1 |
|  | rs200213497 | c.44T->C | V15A | missense | 4 | 1 | 1 | 0 | 4 | 7 | 0 | 69.0 | 41.9 | 22.5 | 62.1 | 13.8 | 0.7 | 0 |
|  | rs2229127 | c.73G->A | V25M | missense | 4 | 0 | 0 | 0 | 3 | 1 | 0 | 1.9 | 166.9 | 15.0 | 22.6 | 12.7 | 0.1 | 0.1 |
|  | rs368827801 | c.125G>A | R42Q | missense | 0 | 0 | 0 | 0 | 0 | 0 | 1 | 0.5 | 0.5 | 0.0 | 5.6 | 19.5 | 0.1 | 0.4 |
|  | rs1397542892 | c.200T>C | L67P | missense | 0 | 0 | 1 | 0 | 1 | 0 | 0 | 0.2 | 0.1 | 3.7 | 5.6 | 31 | 0 | 1 |
|  | rs775137134 | c.319A>G | M107V | missense | 0 | 1 | 0 | 0 | 0 | 0 | 0 | 0.0 | 0.1 | 3.7 | 0.0 | 23.9 | 0 | 0.3 |
|  | rs199969416 | c.409C>T | R137C | missense | 1 | 0 | 0 | 0 | 0 | 0 | 0 | 0.0 | 0.0 | 3.7 | 0.0 | 24.9 | 0.1 | 0.9 |
|  | rs200973220 | c.715G>A | G204S | missense | 0 | 0 | 0 | 0 | 0 | 1 | 0 | 0.2 | 0.1 | 0.0 | 5.6 | 21.9 | 0.2 | 0.2 |
|  | rs4987262 | c.634C->T | R212C | missense | 8 | 1 | 3 | 9 | 6* | 7 | 0 | 111.6 | 67.4 | 78.7 | 73.4 | 27.7 | 0 | 0.9 |
|  | rs2229131 | c.635G->A | R212H | missense | 1 | 0 | 0 | 0 | 0 | 0 | 0 | 0.5 | 9.8 | 3.7 | 0 | 22.3 | 0.1 | 0 |
|  | rs138619017 | c.676C->A | P226T | missense | 9 | 0 | 1 | 1 | 2 | 3 | 0 | 30.5 | 17.4 | 41.2 | 28.2 | 15.3 | 0.2 | 0 |
|  | rs144910793 | c.691G->A | G231R | missense | 1 | 0 | 1 | 0 | 1 | 1 | 0 | 0.5 | 49.9 | 7.5 | 11.3 | 9.1 | 0.4 | 0 |
|  | rs147725404 | c.904G>A | D267N | missense | 0 | 0 | 0 | 0 | 0 | 1 | 0 | 0.0 | 13.9 | 0.0 | 5.6 | 21.6 | 0.2 | 0.1 |
|  | rs28590598 | c.956C->G | S319W | missense | 8 | 0 | 3 | 0 | 4 | 0 | 0 | 5.6 | 282.4 | 41.2 | 22.6 | 9.3 | 0 | 0.7 |
|  | rs141866414 | c.1154C>T | S350L | missense | 0 | 0 | 0 | 0 | 0 | 1 | 0 | 1.1 | 0.8 | 0.0 | 5.6 | 22.1 | 0 | 0 |
|  | rs199793418 | c.1144G>A | A382T | missense | 0 | 0 | 0 | 0 | 1 | 0 | 0 | 0.3 | 0.4 | 0.0 | 5.6 | 22.2 | 0 | 0.8 |

**Table S4:**
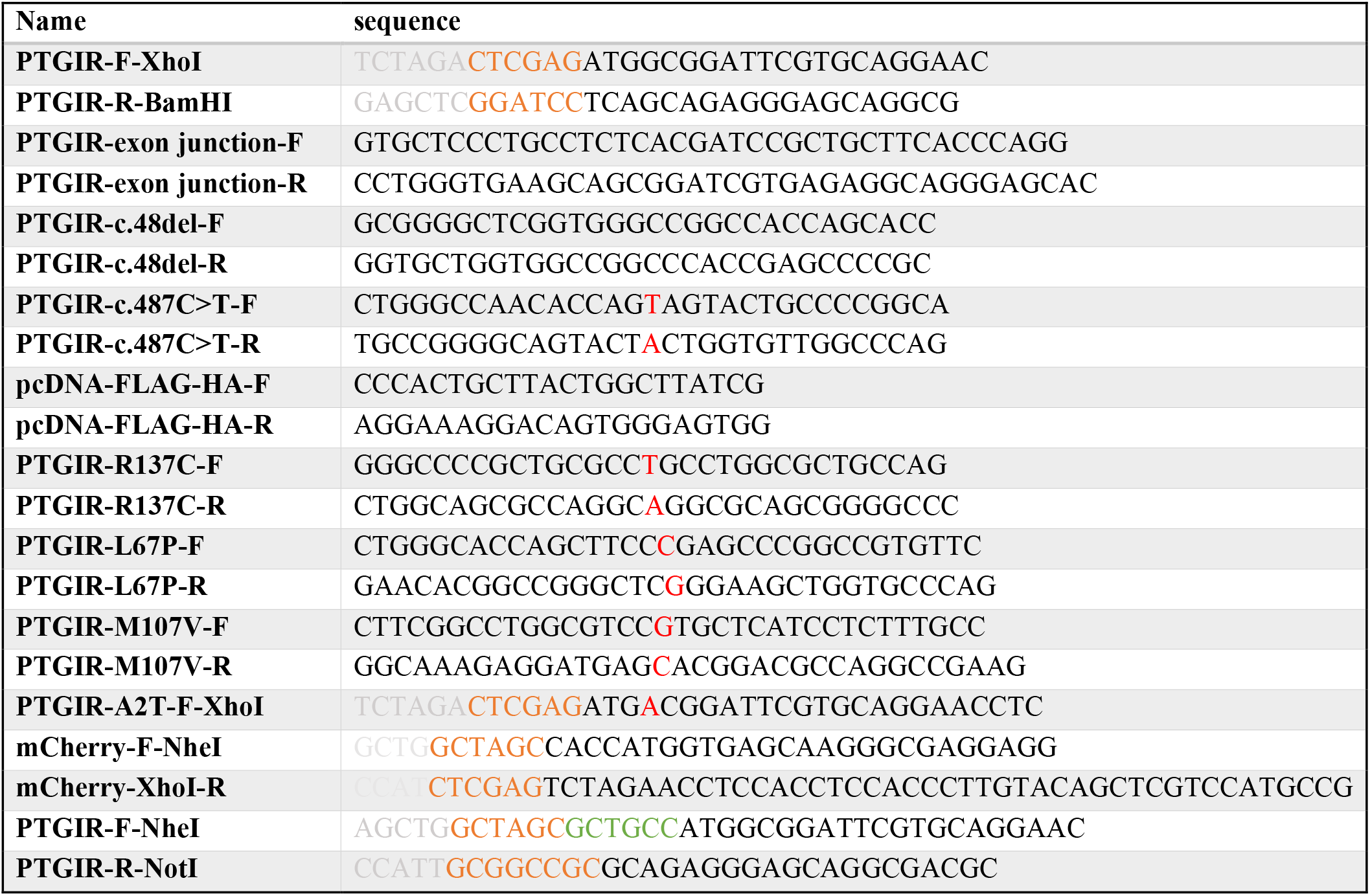
List of PCR primers used in this study. Red : mismatched base to introduce mutation. Orange : Restriction site. Green : Kozak sequence. Grey : spacer (absent of plasmid)

**Table S5:** List of plasmids used in this study.

| **Name** | **Resistance** | **Base vector** | **Cloning method** |
| --- | --- | --- | --- |
| **pcDNA-FLAG-HA** | Ampicillin | Addgene #52535 |  |
| **FLAG-HA-genomicPTGIR** | Ampicillin | pcDNA-FLAG-HA | PTGIR cloned with its intron between XhoI/BamHI sites |
| **FLAG-HA-PTGIR** | Ampicillin | pcDNA-FLAG-HA | coding sequence of PTGIR cloned between XhoI/BamHI sites |
| **FLAG-HA-PTGIR-c.48del** | Ampicillin | pcDNA-FLAG-HA | coding sequence of PTGIR cloned between XhoI/BamHI sites, c.48del mutant (rs754755149) |
| **FLAG-HA-PTGIR-c.487C>T** | Ampicillin | pcDNA-FLAG-HA | coding sequence of PTGIR cloned between XhoI/BamHI sites, c.487C->T mutant (rs199560500) |
| **FLAG-HA-PTGIR-R137C** | Ampicillin | pcDNA-FLAG-HA | coding sequence of PTGIR cloned between XhoI/BamHI sites, R137C mutant |
| **pcDNA-mCherry** | Ampicillin | pcDNA-FLAG-HA | coding sequence of mCherry cloned between NheI/XhoI sites, replacing FLAG-HA tag |
| **FLAG-HA-PTGIR-L67P** | Ampicillin | pcDNA-FLAG-HA | coding sequence of PTGIR cloned between XhoI/BamHI sites, L67P mutant |
| **FLAG-HA-PTGIR-M107V** | Ampicillin | pcDNA-FLAG-HA | coding sequence of PTGIR cloned between XhoI/BamHI sites M107V mutant |
| **FLAG-HA-PTGIR-A2T** | Ampicillin | pcDNA-FLAG-HA | coding sequence of PTGIR cloned between XhoI/BamHI sites A2T mutant |
| **pcDNA-PCOLCE2** | Ampicillin | pcDNA-FLAG-HA | Insertion of PCOLCE2 (no stop codon) NheI/NotI sites (PCR), removes FLAG-HA tag |
| **pcDNA-PCOLCE2-mCherry-HA** | Ampicillin | pcDNA-PCOLCE2 | Insertion of mCherry NotI/EcoRI sites (PCR) |
| **pcDNA-PTGIR-mCherry** | Ampicillin | pcDNA-PCOLCE2-mCherry-HA | Insertion of PTGIR sequence (without stop codon) between NheI/NotI sites of PCOLCE2-mCherry, replacing PCOLCE2 |

## Supplementary Figure Legends

**Figure S1:**
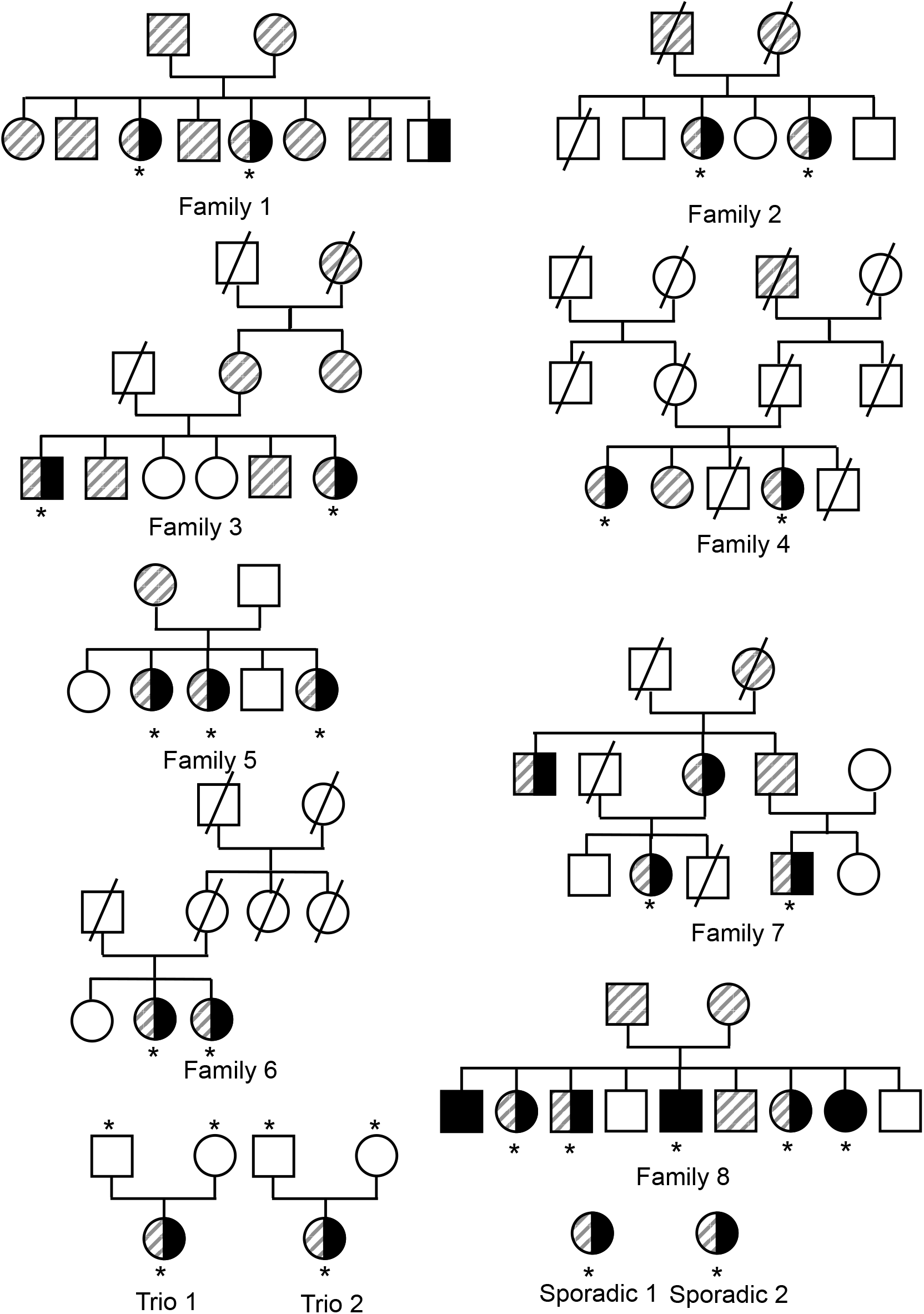
Families trees of FMD families. * indicates the subjects for which exome-sequencing was performed. Black color indicates a confirmed FMD diagnosis. Hatching indicates subjects diagnosed with hypertension.

**Figure S2:**
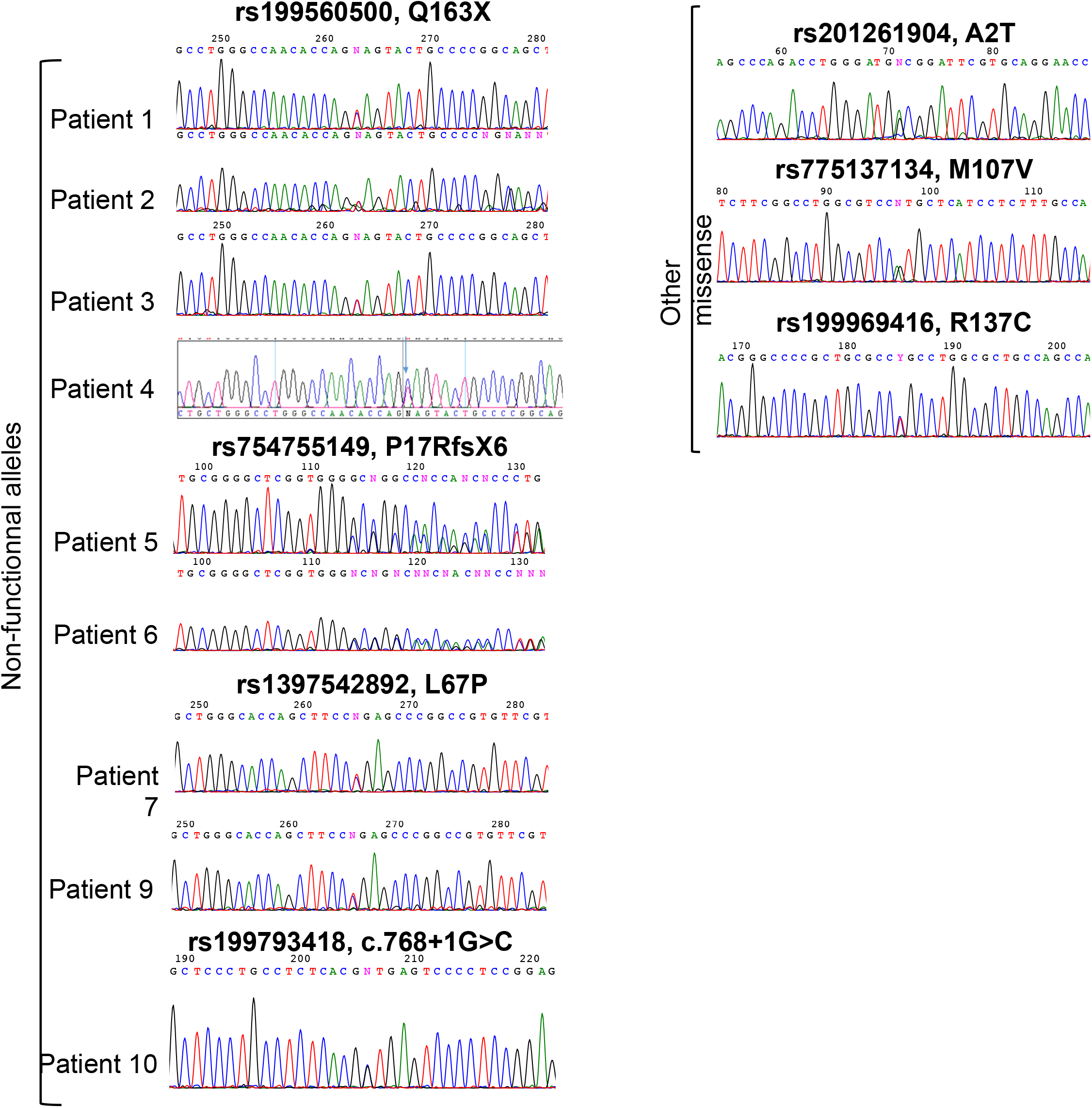
Chromatograms showing the nonsense and uncharacterized missense variants identified in FMD patients.

**Figure S3:**
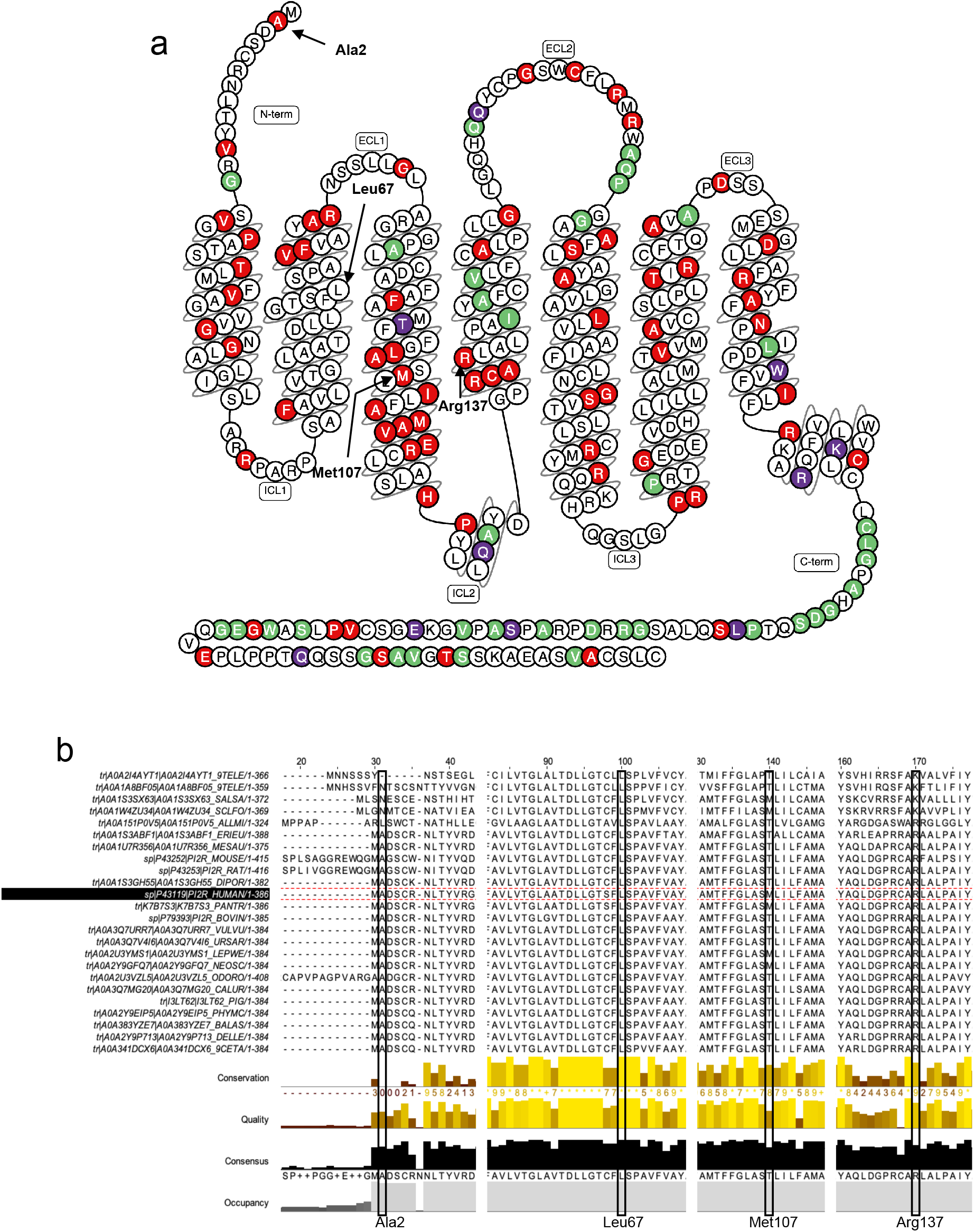
Position and conservation of newly characterized IP mutants. a) Representation of IP protein secondary structure, with its 7 transmembrane domains and N- and C-terminal domains. Colored residues indicate possible deleterious mutations due to rare or common variants, based on SIFT/PolyPhen scores (red/purple = deleterious, green = tolerated). Black arrows indicate the four previously uncharacterized mutants identified in this study. b) Alignment of human IP with orthologs in vertebrates. Prostacyclin receptor sequences were recovered from Uniprot, keeping only full-length IP proteins in vertebrates. Sequences were aligned using Clustal Omega on EBI webserver (www.ebi.ac.ul/Tools/msa.clustalo) and alignments were visualized using Jalview (www.jalview.org).

**Figure S4:**
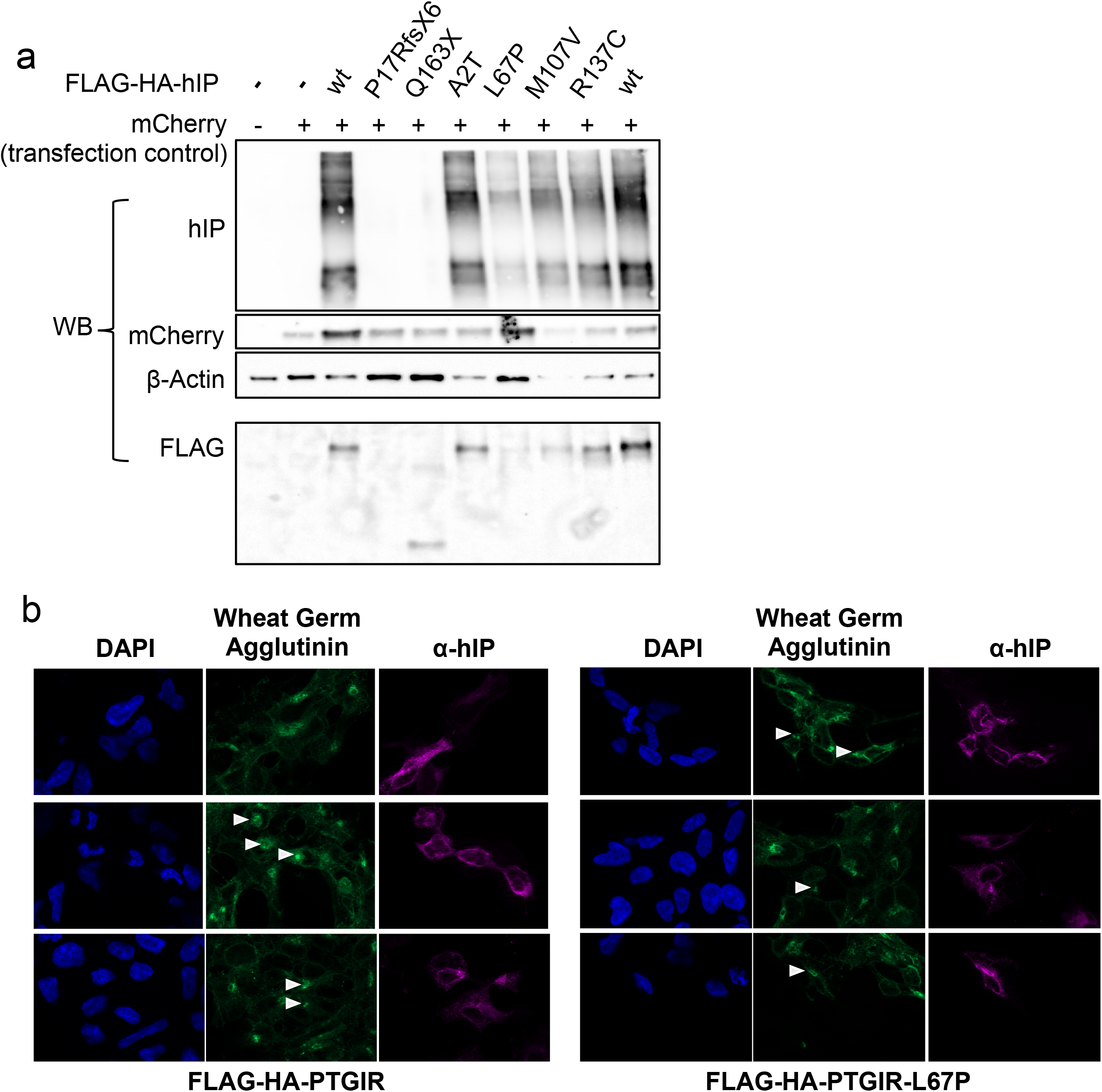
Evaluation of mutant IP protein expression and localization. a) SDS-PAGE/Western Blot assay on whole cells extracts of HEK293 cells overexpressing wild-type or mutant IP (with FLAG-HA N-terminal tag) and mCherry. mCherry was used as a transfection control to evaluate the success and homogeneity of transfection in each sample. hIP, mCherry and β-Actin were detected using specific primary antibodies. hIP is detected as a mix of bands and higher molecular weight smear due to its extensive post-translational modification. This independent experiment (compared to Figure 3) shows the expression of L67 hIP mutant with a similar appearance compared to WT hIP, although at a lower level. b) Immunofluorescence visualization of HEK293 cells overexpressing wild-type or L67P hIP. hIP localization was assayed using hIP specific antibody and Alexa647-conjugated secondary antibody (purple signal). Fixed cells were incubated with Alexa488-conjugated Wheat Germ Agglutinin (WGA, green signal), recognizing phosphoproteins, and with DAPI to stain DNA (blue signal). Images were taken with 100x objective on a Zeiss ApoTome system. White arrows show the position of intracellular aggregates stained by WGA, corresponding to the endoplasmic reticulum/Golgi apparatus, where hIP staining is clearly visible on L67P mutant, but not on WT protein.

**Figure S5:**
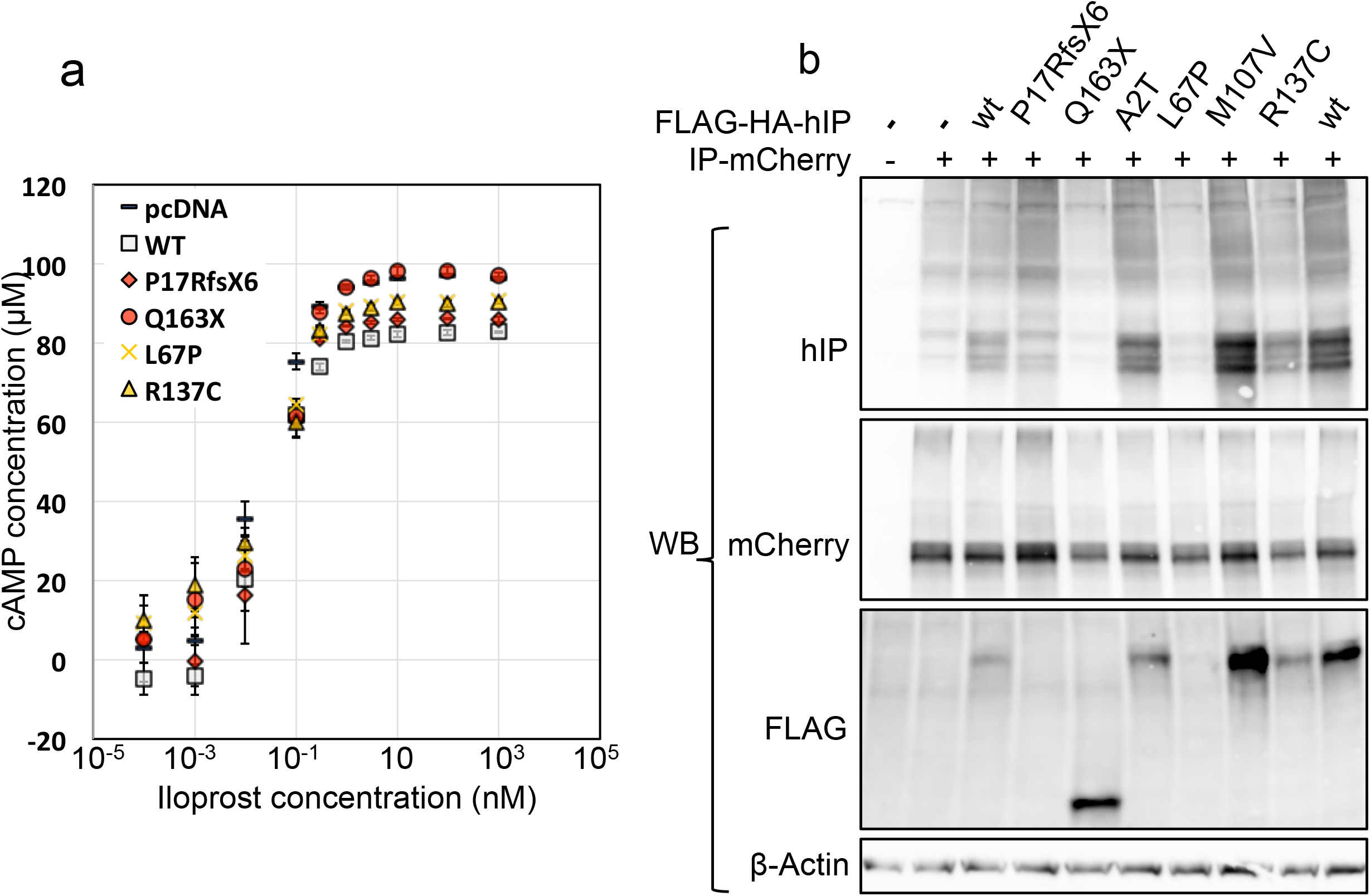
Evaluation of mutant IP effect on co-expressed wild-type IP. a) Concentration of cAMP in HEK293 cells overexpressing wild-type hIP-mCherry and wild-type or mutant FLAG-HA-hIP or pcDNA-FLAG-HA (pcDNA). HEK293 cells were treated with various doses of Iloprost for 25 minutes, before cAMP concentration was measured using cAMP-Glo assay. Error bars represent the standard deviation of three biological replicates. b) SDS-PAGE/Western Blot assay on whole cells extracts of HEK293 cells overexpressing wild-type or mutant hIP (with FLAG-HA N-terminal tag) and wild-type IP-mCherry. mCherry antibody was used to detect crude IP-mCherry, FLAG antibody to detect FLAG-HA-IP, and hIP antibody to detect both hIP and its more modified forms (including N-terminally cleaved, isoprenylated and palmitoylated forms).

**Figure S6:**
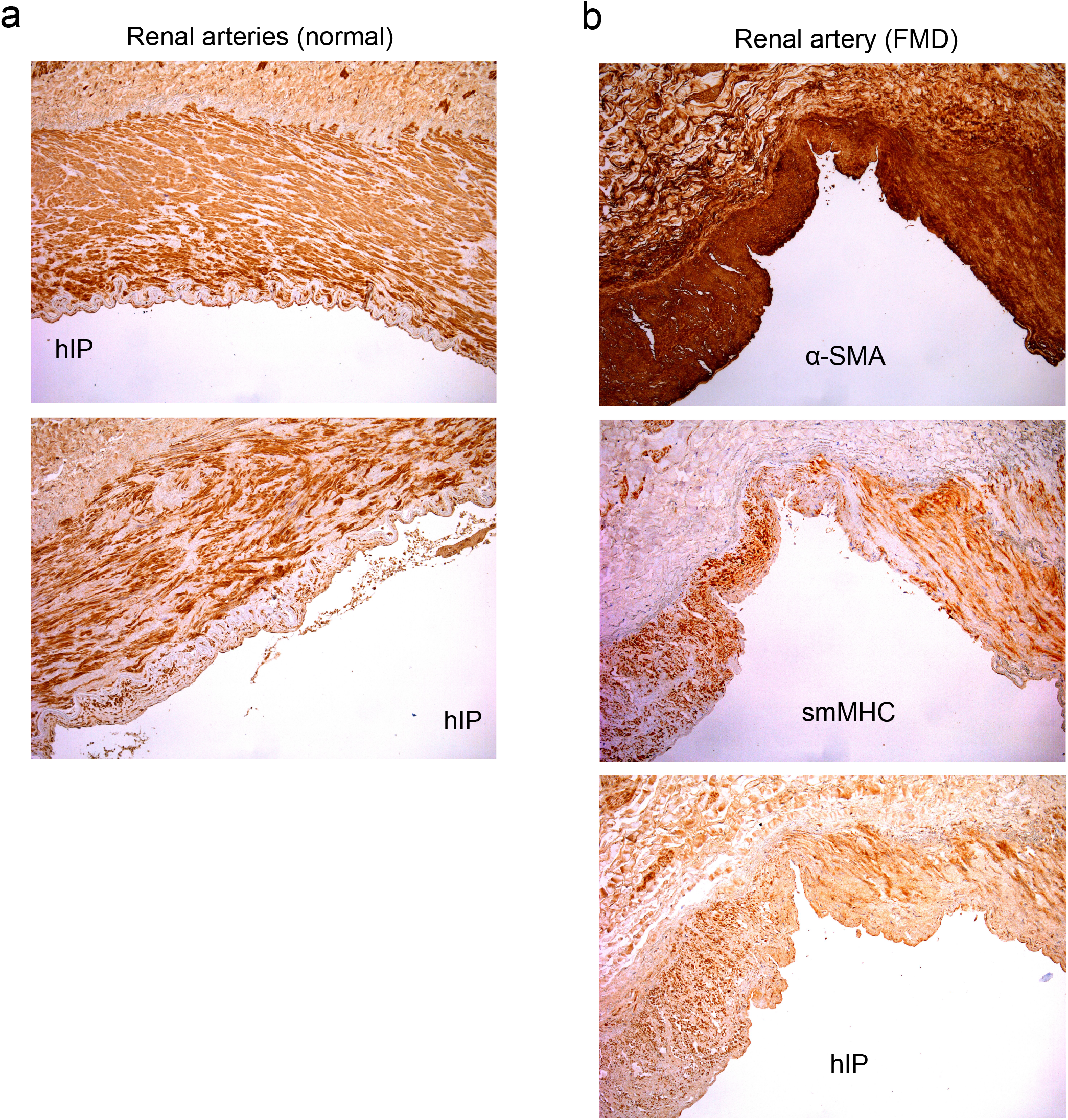
Immunohistochemical visualization of IP and smooth muscle cells markers in normal arteries or in an FMD lesion. Specific antibodies targeting human prostacyclin receptor (hIP), smooth muscle Actin (α-SMA) and smooth muscle Myosin heavy chain (smMHC) were used to detect these proteins in 2 normal renal arteries (a) and one renal artery with FMD lesion close to an aneurysm (b). Positive labeling is indicated by orange/brown color.

## Bibliography

1. Gornik HL, Persu A, Adlam D et al. First International Consensus on the diagnosis and management of fibromuscular dysplasia. Vascular medicine (London, England) 2019;24:164–189.

2. Olin JW, Froehlich J, Gu X et al. The United States Registry for Fibromuscular Dysplasia: results in the first 447 patients. Circulation 2012;125:3182–90.

3. Stanley JC, Gewertz BL, Bove EL, Sottiurai V, Fry WJ. Arterial fibrodysplasia. Histopathologic character and current etiologic concepts. Archives of surgery (Chicago, Ill : 1960) 1975;110:561–6.

4. Luscher TF, Lie JT, Stanson AW, Houser OW, Hollier LH, Sheps SG. Arterial fibromuscular dysplasia. Mayo Clinic proceedings 1987;62:931–52.

5. Di Monaco S, Georges A, Lengele JP, Vikkula M, Persu A. Genomics of Fibromuscular Dysplasia. International journal of molecular sciences 2018;19.

6. Miller DJ, Marin H, Aho T, Schultz L, Katramados A, Mitsias P. Fibromuscular dysplasia unraveled: the pulsation-induced microtrauma and reactive hyperplasia theory. Medical hypotheses 2014;83:21–4.

7. Sottiurai V, Fry WJ, Stanley JC. Ultrastructural characteristics of experimental arterial medial fibroplasia induced by vasa vasorum occlusion. The Journal of surgical research 1978;24:167–77.

8. Hayes SN, Kim ESH, Saw J et al. Spontaneous Coronary Artery Dissection: Current State of the Science: A Scientific Statement From the American Heart Association. Circulation 2018;137:e523–e557.

9. Adlam D, Alfonso F, Maas A, Vrints C. European Society of Cardiology, acute cardiovascular care association, SCAD study group: a position paper on spontaneous coronary artery dissection. European heart journal 2018;39:3353–3368.

10. Olin JW, Di Narzo AF, d’Escamard V et al. A Plasma Proteogenomic Signature for Fibromuscular Dysplasia. Cardiovascular research 2019.

11. Kiando SR, Tucker NR, Castro-Vega LJ et al. PHACTR1 Is a Genetic Susceptibility Locus for Fibromuscular Dysplasia Supporting Its Complex Genetic Pattern of Inheritance. PLoS genetics 2016;12:e1006367.

12. Adlam D, Olson TM, Combaret N et al. Association of the PHACTR1/EDN1 Genetic Locus With Spontaneous Coronary Artery Dissection. Journal of the American College of Cardiology 2019;73:58–66.

13. Marian AJ. Molecular genetic studies of complex phenotypes. Translational research : the journal of laboratory and clinical medicine 2012;159:64–79.

14. Dobrowolski P, Januszewicz M, Klisiewicz A et al. Echocardiographic assessment of left ventricular morphology and function in patients with fibromuscular dysplasia: the ARCADIA-POL study. Journal of hypertension 2018;36:1318–1325.

15. Kiando SR, Barlassina C, Cusi D et al. Exome sequencing in seven families and gene-based association studies indicate genetic heterogeneity and suggest possible candidates for fibromuscular dysplasia. Journal of hypertension 2015;33:1802-10; discussion 1810.

16. Guo MH, Plummer L, Chan YM, Hirschhorn JN, Lippincott MF. Burden Testing of Rare Variants Identified through Exome Sequencing via Publicly Available Control Data. American journal of human genetics 2018;103:522–534.

17. Huang da W, Sherman BT, Lempicki RA. Systematic and integrative analysis of large gene lists using DAVID bioinformatics resources. Nature protocols 2009;4:44–57.

18. Szklarczyk D, Gable AL, Lyon D et al. STRING v11: protein-protein association networks with increased coverage, supporting functional discovery in genome-wide experimental datasets. Nucleic acids research 2019;47:D607–d613.

19. Tranchevent LC, Ardeshirdavani A, ElShal S et al. Candidate gene prioritization with Endeavour. Nucleic acids research 2016;44:W117–21.

20. Majed BH, Khalil RA. Molecular mechanisms regulating the vascular prostacyclin pathways and their adaptation during pregnancy and in the newborn. Pharmacological reviews 2012;64:540–82.

21. Turley TN, Theis JL, Sundsbak RS et al. Rare Missense Variants in TLN1 Are Associated With Familial and Sporadic Spontaneous Coronary Artery Dissection. Circulation Genomic and precision medicine 2019;12:e002437.

22. Khera AV, Won HH, Peloso GM et al. Diagnostic Yield and Clinical Utility of Sequencing Familial Hypercholesterolemia Genes in Patients With Severe Hypercholesterolemia. Journal of the American College of Cardiology 2016;67:2578–89.

23. Arehart E, Stitham J, Asselbergs FW et al. Acceleration of cardiovascular disease by a dysfunctional prostacyclin receptor mutation: potential implications for cyclooxygenase-2 inhibition. Circulation research 2008;102:986–93.

24. Stitham J, Arehart E, Elderon L et al. Comprehensive biochemical analysis of rare prostacyclin receptor variants: study of association of signaling with coronary artery obstruction. The Journal of biological chemistry 2011;286:7060–9.

25. Insel PA, Murray F, Yokoyama U et al. cAMP and Epac in the regulation of tissue fibrosis. British journal of pharmacology 2012;166:447–56.

## References

1. Saw J, Aymong E, Sedlak T et al. Spontaneous coronary artery dissection: association with predisposing arteriopathies and precipitating stressors and cardiovascular outcomes. Circulation Cardiovascular interventions 2014;7:645–55.

2. Tweet MS, Eleid MF, Best PJ et al. Spontaneous coronary artery dissection: revascularization versus conservative therapy. Circulation Cardiovascular interventions 2014;7:777–86.

3. Tang WH, Hazen SL. The contributory role of gut microbiota in cardiovascular disease. The Journal of clinical investigation 2014;124:4204–11.

4. Huang da W, Sherman BT, Lempicki RA. Systematic and integrative analysis of large gene lists using DAVID bioinformatics resources. Nature protocols 2009;4:44–57.

5. Szklarczyk D, Gable AL, Lyon D et al. STRING v11: protein-protein association networks with increased coverage, supporting functional discovery in genome-wide experimental datasets. Nucleic acids research 2019;47:D607–d613.

6. Tranchevent LC, Ardeshirdavani A, ElShal S et al. Candidate gene prioritization with Endeavour. Nucleic acids research 2016;44:W117–21.

7. Guo MH, Plummer L, Chan YM, Hirschhorn JN, Lippincott MF. Burden Testing of Rare Variants Identified through Exome Sequencing via Publicly Available Control Data. American journal of human genetics 2018;103:522–534.

8. Horn M, Geisen C, Cermak L et al. DRE-1/FBXO11-dependent degradation of BLMP-1/BLIMP-1 governs C. elegans developmental timing and maturation. Developmental cell 2014;28:697–710.

9. Kiando SR, Barlassina C, Cusi D et al. Exome sequencing in seven families and gene-based association studies indicate genetic heterogeneity and suggest possible candidates for fibromuscular dysplasia. Journal of hypertension 2015;33:1802-10; discussion 1810.

